# CLB101^TM^ Improves Gastrointestinal Symptom Burden in Overweight Adults: A Randomized, Double-Blind, Placebo-Controlled Study

**DOI:** 10.64898/2026.09.03.26362186

**Authors:** Azure D. Grant, Brian Meehan, Marie Crisel E. Krupa, Miguel A. Rosales, Karol Sokolowski, Brett Newswanger, David Keller, Veronica Luna, Erin Marie S. Valentin, Thomas McCormick, Mahmoud Ghannoum, Noah Craft

**Affiliations:** People Science, Inc, Los Angeles, CA; ClostraBio, Inc., Chicago, IL, USA; Keller Consulting Group, Beachwood, Ohio, USA; Case Western Reserve University School of Medicine, Cleveland, Ohio, USA; Fungal Discovery Research, Cleveland, Ohio, USA; University Hospitals Cleveland Medical Center, Cleveland, Ohio, USA

## Abstract

**Background:** *Anaerostipes caccae* CLB101^TM^ is a next-generation probiotic that directly produces butyrate and is hypothesized to reduce gastrointestinal symptoms by strengthening intestinal barrier integrity. Clinical evidence for *A. caccae* in symptomatic adult populations remains limited.

**Methods:** Ninety-three adults were randomized 1:1 to CLB101^TM^ (n=47) or placebo (n=46) for 28 days. The primary outcome was safety and tolerability as assessed by the number, frequency, and severity of adverse events. Secondary outcomes were focused on exploring signals of efficacy, and included change in the Gastrointestinal Symptom Rating Scale (GSRS) composite score from baseline to Week 4. Five GSRS subcategories were additionally evaluated. Exploratory endpoints included daily self-reported GI symptoms, as well as stool and blood measures of intestinal barrier function.

**Results:** 71 participants were evaluated for the GSRS (77.5% female, aged 42.1 SD 12 years, BMI mean 31.3 SD 3.6). No adverse events were reported in the placebo arm, while three self-limiting adverse events were associated with CLB101^TM^ administration. CLB101^TM^ reduced GI symptom burden versus placebo on the primary GSRS composite at Week 4 (mean change: −1.60 vs −0.90; MWU p=0.040). Two secondary subscales reached significance after FDR-BH correction: Reflux (mean change: −1.26 vs −0.30; FDR-adjusted p=0.024) and Abdominal Pain (mean change: −1.86 vs −0.83; FDR-adjusted p=0.024). Daily symptom diaries showed trends toward greater reductions in abdominal discomfort, bloating, and flatulence in the active arm at the end of study. This study population did not demonstrate signs of intestinal barrier dysfunction. Product adherence averaged 97.7%.

**Conclusions:** CLB101^TM^ was safe and tolerable and significantly improved overall GI symptom burden, reflux, and abdominal pain after four weeks of daily supplementation in overweight symptomatic adults.

**Trial Registration:** The trial was approved by Sterling IRB (14583-NACraft) and registered with ClinicalTrials.gov (NCT07336615).

## INTRODUCTION

*Anaerostipes caccae* CLB101™ is a next-generation probiotic isolated from the gut of a healthy infant, administration of which is intended to increase intestinal butyrate production. Butyrate is a naturally occurring small molecule produced by a small number of bacteria found in the gut of healthy individuals, where it serves as the primary fuel source for colonocytes, helps maintain epithelial barrier integrity via tight-junction assembly and mucus production, and reduces intestinal inflammation (Hamer et al., 2008; Peng et al., 2009; Liu et al., 2018; Recharla et al., 2023; Dagbasi et al., 2024).

*A. caccae* was first described (Schwiertz et al., 2002) as an obligately anaerobic, saccharolytic bacterium isolated from human feces that produces acetate, butyrate, and lactate from glucose metabolism. It belongs to *Clostridium* cluster XIVa and is recognized as one of the key butyrate-producing genera in the human gut, alongside *Faecalibacterium*, *Roseburia*, and *Coprococcus* (Sola et al., 2026). In addition to producing butyrate from certain monosaccharides and prebiotics, *A. caccae* was shown to convert lactate and acetate into butyrate (Duncan et al., 2004), thus serving as the terminal node in cross-feeding networks that may include ubiquitous commensals like *Bifidobacteria* and *Bacteroides* (Belenguer et al., 2006; Falony et al., 2016; Chia et al., 2020a). Bacterial production of butyrate in turn drives an upregulation of intestinal mucus production via the MUC2 gene, which provides a growth substrate for *Akkermanisa muciniphila* and other microbes associated with the mucosal layer (Chia et al., 2018).The central role played by *A. caccae* in these trophic interactions suggests it is a keystone species in intestinal health.

To that end, reduced abundance of the *Anaerostipes* genus has been reported in individuals with irritable bowel syndrome (IBS), inflammatory bowel disease (IBD), and colorectal cancer (Lo Presti et al., 2019; Ryan et al., 2020; Song et al., 2024). In addition, studies of food allergic individuals have shown that *A. caccae* is significantly reduced in abundance compared to healthy individuals (Feehley et al., 2019). In preclinical models, *A. caccae* and lactulose synbiotic treatment prevents and treats food allergy (Hesser et al., 2024) and reduces intestinal inflammation (Qiao & Gao, 2026) while reducing uric acid. Furthermore, *A. caccae* has been shown to treat sorbitol intolerance (Lee et al., 2024), further supporting its role in reducing reactivity to problematic foods.

*A. caccae* displays promising properties for promoting healthy gut physiology. Moreover, as a feasible probiotic supplement, in comparison to the high-oxygen sensitivity of most butyrate producers, *A. caccae* spores survived 15 weeks under atmospheric oxygen (Kadowaki et al., 2023). Additionally, co-administration of *A. caccae* strain L2 with galacto-oligosaccharides (a synbiotic approach) significantly enhances cecal and fecal butyrate concentrations compared to controls (Sato et al., 2008), and may be an early beneficial colonizer of the infant gut (Chia et al., 2020b; 2021).

In humans, barrier dysfunction is well documented in intestinal disease states such as IBD, IBS, and celiac disease, but its role in otherwise healthy populations is less clear (Camilleri, 2019; Hoshiko et al., 2021). Colloquially, the term “leaky gut” has been used to refer to a collection of symptoms that may or may not be associated with adverse intestinal absorption of various molecules via the paracellular route. Biomarkers used to quantify intestinal permeability are varied, and include the urinary excretion of lactulose/mannitol (or other sugars), serum lipopolysaccharide binding protein (LBP), as well as fecal levels of calprotectin and zonulin (Camilleri, 2019). Each of these biomarkers has its challenges, but collectively may represent a comprehensive approach to characterizing potential barrier dysfunction. Overweight and obese individuals have been reported to show increased intestinal permeability (Teixeira et al., 2012), though results have been mixed (Brignardello et al., 2010; Damms-Machado et al., 2017). In an effort to interrogate the impact of CLB101^TM^ on intestinal barrier function and GI symptoms, we enrolled overweight individuals with elevated symptomatic GI distress scores as measured by the clinically-validated Gastrointestinal System Rating Scale (GSRS) (Talley et al., 2001). Together, this randomized, placebo-controlled, double-blind study investigated the safety and efficacy of CLB101™ on digestive symptom alleviation, reduction of inflammatory markers, improvement of gut barrier integrity and impact on microbiome composition in overweight symptomatic adults.

## Methods

### Eligibility

Eligible participants were healthy American adults aged 18–65 years with a BMI of 27-35 kg/m², who reported moderate to severe gastrointestinal symptoms (e.g., abdominal pain, bloating, flatulence, constipation, or diarrhea) and had a moderate GSRS score (4-7). Individuals using medications for anxiety, cannabis, or nicotine were required to be on stable regimens for at least 4 weeks prior to randomization and throughout the study.

Key exclusion criteria included use of antibiotics, probiotics, or prebiotics within 4 weeks prior to randomization or during the study; or regular use of medications known to impact gastrointestinal function or the microbiome (e.g., immunosuppressants, systemic steroids, antifungals, NSAIDs). Individuals following restrictive diets (e.g., carnivore, raw, fruitarian, or liquid diets), consuming more than two alcoholic drinks per day, weekly or more frequent cannabis or nicotine use, or any substance use disorder were excluded. Participants with clinically diagnosed gastrointestinal diseases (including but not limited to inflammatory bowel disease, gastroesophageal reflux disease, ulcers, celiac disease, diverticular disease, pancreatitis, gastroparesis, gallbladder disease, or gastrointestinal cancers), significant liver disease (NAFLD was not an exclusion criterion), gastrointestinal infections (e.g., *Clostridium difficile*, *Helicobacter pylori*, parasitic infections), prior gastrointestinal surgery (excluding appendectomy or cholecystectomy), or history of gastrointestinal bleeding or perforation were excluded. Pregnant or breastfeeding women were excluded.

### Study Design

Participants completed up to an 8-week study consisting of a 2-week Randomization and Shipping Period, a 2-week Baseline Period, and a 4-week Product/Placebo Use Period. Demographic and medical history data were collected, including concomitant medications, during the screening period. Participants were randomized to one of two groups: (1) CLB101™ Probiotic Supplement, and (2) matching placebo. The investigators, the study team and participants were blinded to the CLB101^TM^ assignment. This study was conducted remotely using the web-based data collection platform (described below) called Consumer Health Learning and Organizing Ecosystem (Chloe) by People Science (People Science Inc, Los Angeles, CA). Participants completed study assessments using the Chloe app during the course of the study. Participants’ activities are described in **(Table 1 Study Timeline).**

**Table 1.**
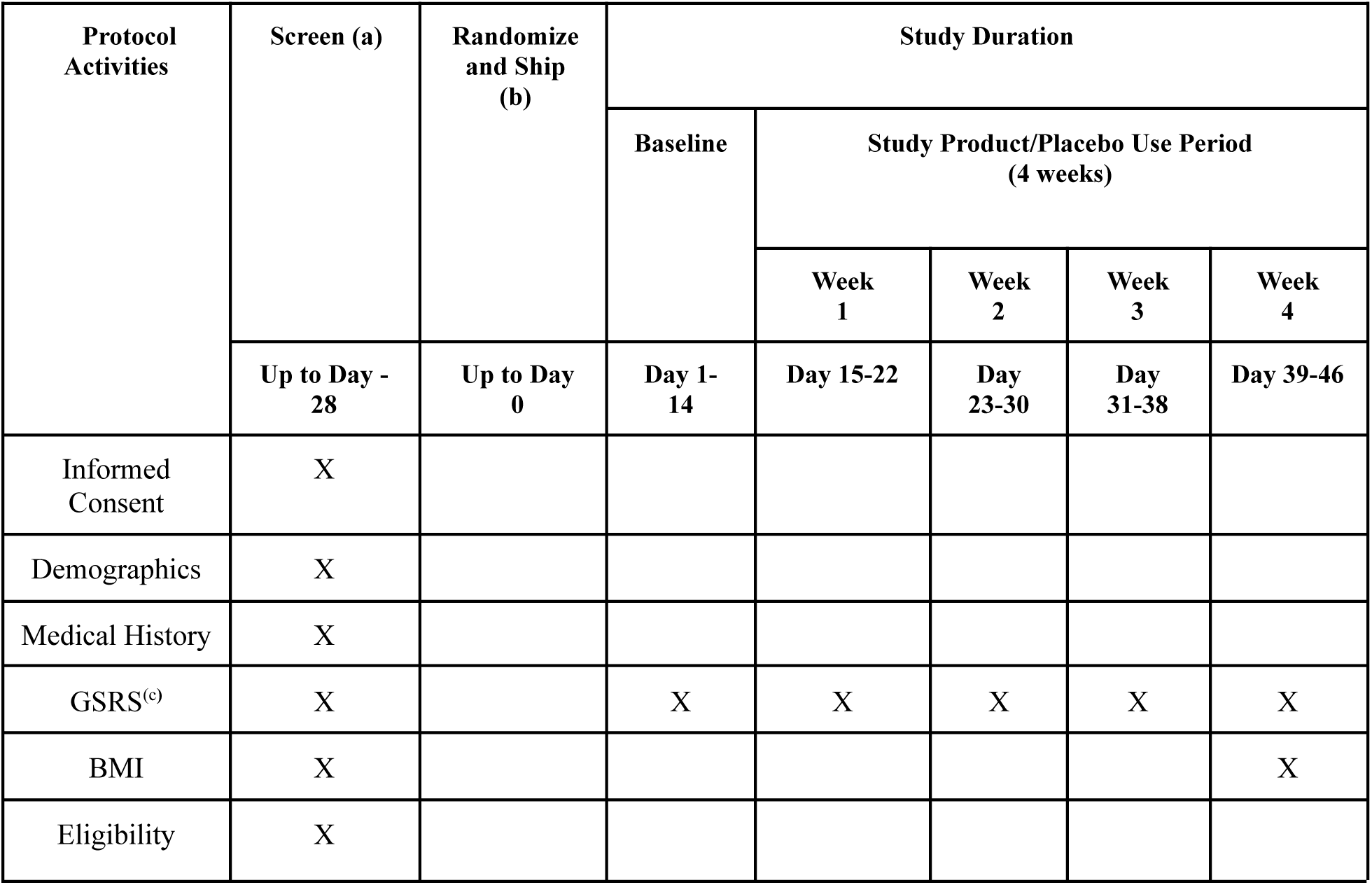
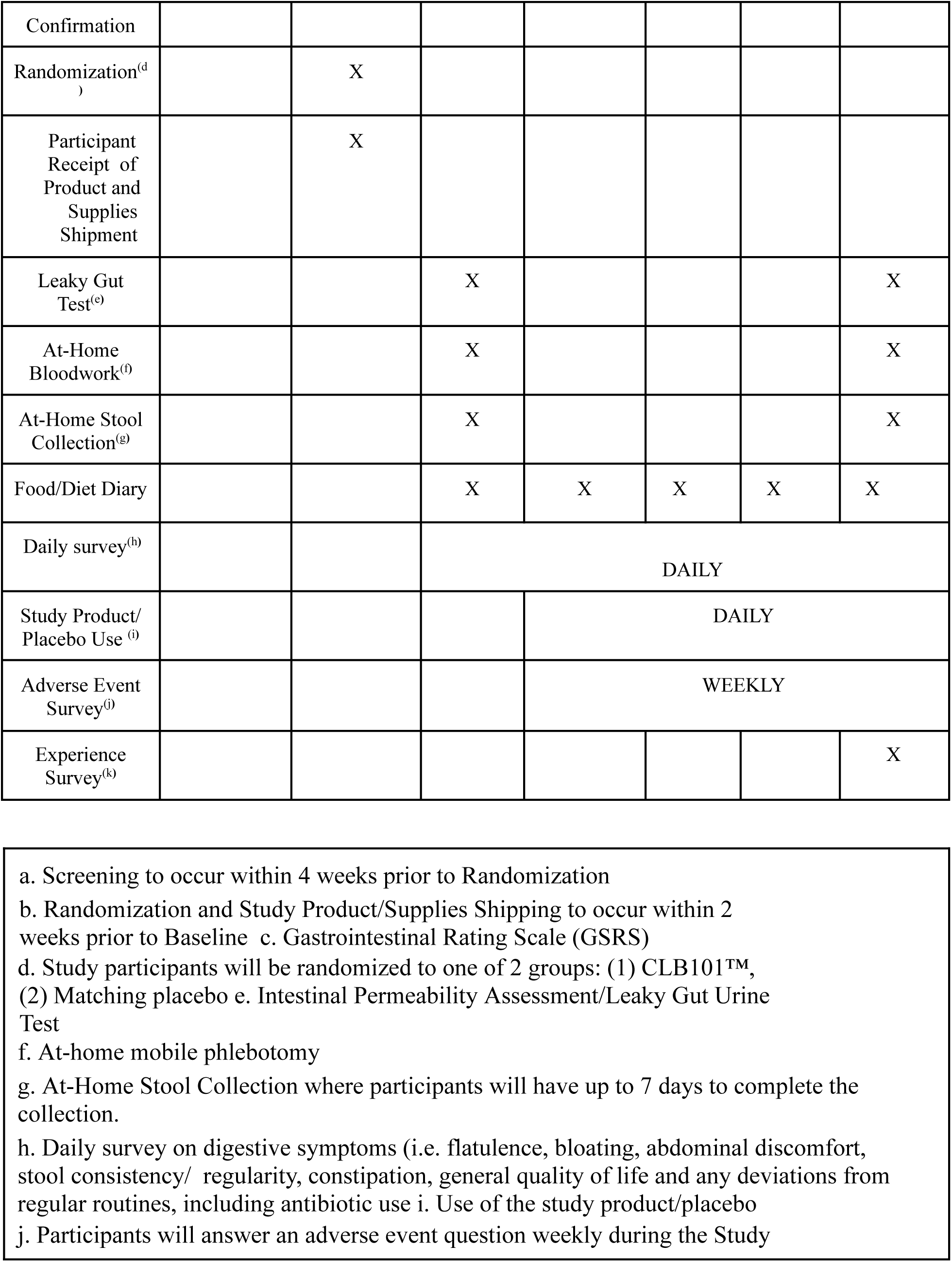
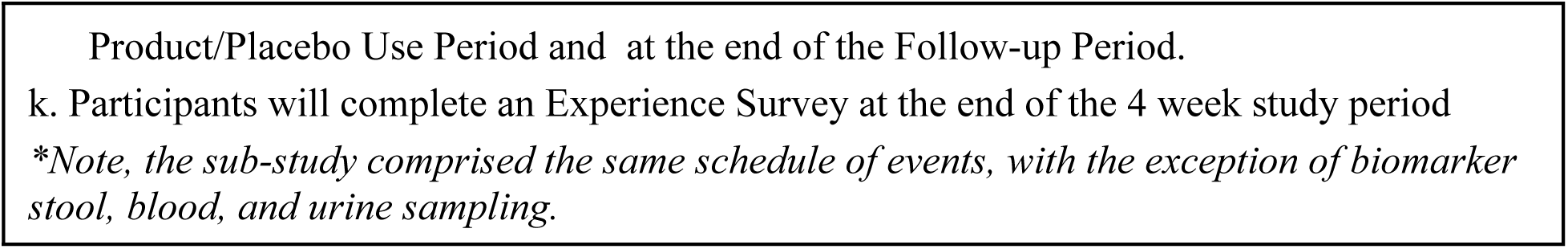
Study Activities.

### Data Management System

All data were securely stored on Chloe Amazon Web Services HIPAA-compliant servers. The Chloe platform contains modules for building and managing surveys, study landing pages, marketing outreach with tracking tools for recruitment, audited electronic consent forms, data management and analytics using an integrated relational database. Additionally, the platform contains a user-facing app for participants that delivers surveys, study instructions, calendar reminders, communication with study team, and personal data reports at study culmination. Data from completed assessments were automatically collected for analysis.

### Recruitment, Consent, and Enrollment

Participants were recruited through social media channels and researcher networks. Recruitment outreach consisted of IRB-approved advertising by email, digital marketing channels and word of mouth. The IRB-approved study landing page hosted on the People Science website led to an IRB-approved pre-screening questionnaire to determine individual qualification. Qualified participants were invited to download the Chloe application and create an account to access the Informed Consent Form and continue through eligibility determination and enrollment.

Virtual electronic informed consent, including a study specific privacy authorization and the California Experimental Subject’s Bill of Rights (as applicable) were provided through the HIPAA-compliant cloud-based platform Chloe. Eligible participants who provided virtual electronic consent were automatically registered into the study by the platform.

### Intervention Preparation and Dosing

The probiotic strain *Anaerostipes caccae* CLB101™ a next-generation butyrate-producing probiotic, was selected based on data from pre-clinical models demonstrating positive impacts on intestinal barrier function and immune modulation following exposure to problematic foods. Isolated from a healthy human donor, CLB101™ was manufactured in a GMP-certified facility under stringent quality control standards and achieved self-affirmed GRAS (Generally Recognized as Safe) status following rigorous review of safety data by an independent expert panel, including multiple toxicology studies demonstrating safety and tolerability at doses exceeding human consumption levels (Modica et al., 2025). The dose of 1 billion AFU (Active Fluorescent Units) per day was selected based on preclinical efficacy studies (Hesser et al., 2024). Consistent with other next-generation probiotics, AFU was used to define the dose due to its ability to predict the metabolic activity of viable but not culturable cells (VBNC), which can potentially have a significant impact on human health and are not accounted for when using the traditional enumeration of colony-forming units (CFU) (Visciglia 2022), which in this study product amounted to 10^8^ CFU per dose.

### Blood Collection

ExamOne (Quest Diagnostics, Lenexa, KS) was contracted to provide mobile phlebotomy services for participant safety blood parameters. Fungal Disease Research Organization (FDR) (Cleveland, OH), supplied serum kits directly to People Science. The safety and exploratory efficacy blood parameters collected included serum lipopolysaccharide binding protein (LPS-BP) and ALT/AST at baseline and once at the end of the study. ExamOne mobile phlebotomists were required to ship participant samples back to FDR for analysis. Analyte extraction methods are detailed in the **Supplemental Methods.**

### At-Home Stool Collection and qPCR

FDR at-home stool sample kits included a specimen pan, a 30ml conical bottom container for stool, an attached screwcap with spoon, swabs, and a biological substance shipper. Participants were asked to provide two stool samples throughout the course of the study. One sample was collected during the baseline period, and the second sample was collected at the end of the study. Participants were asked to freeze their stool samples overnight and ship with the swab sample on ice packs the next day at the aforementioned two timepoints. These were analyzed for calprotectin, zonulin, lactoferrin, butyrate, microbiome analysis and qPCR.

Abundance of the target organism, *Anaerostipes caccae*, was quantified by quantitative PCR (qPCR) on DNA extracted from stool swabs. Reactions were carried out in a total volume of 20 µL containing 1× IDT PrimeTime Gene Expression Master Mix (Integrated DNA Technologies, Coralville, Iowa) (10 µL of 2× master mix), 250 nM each of the forward primer (5′-GTTTTCGGATGGATTTCCTATAT-3′) and reverse primer (5′-CTTTTCACACTGAATCATGCGATT-3′), 125 nM dual-labeled hydrolysis probe (5′-/56-FAM/TGGAAACGG/ZEN/CTGCTAATACCGCAT/3IABkFQ/-3′), 2 µL of template DNA, and nuclease-free water. Amplification was performed on an Analytik Jena qTower³ (Analytik Jena, Jena, Germany) system with an initial polymerase activation step at 95 °C for 3 min, followed by 45 cycles of denaturation at 95 °C for 5 s and combined annealing/extension at 62 °C for 30 s. Absolute quantification was performed against a standard curve generated from a serial dilution of a positive-control template of known copy number. Samples that failed to amplify or fell below the limit of detection (LOD) were assigned a value of half the LOD prior to analysis. Other fecal analysis methods are detailed in the **Supplemental Methods**

### Intestinal Permeability Assessment

The Genova Diagnostics Intestinal Permeability Assessment, also known as the Leaky Gut Urine Test (Genova Diagnostics, Asheville, NC) assessed whether the intestinal barrier exhibited increased intestinal permeability. This assessment was completed during baseline and end of study.

### Subjective Daily Assessments

Participants completed daily numeric rating scales (1-10) of abdominal discomfort, bloating, flatulence, and quality of life. Participants also reported daily adherence to product use, any changes to daily medications, as well as consumption of alcohol or evening caffeine.

### Adverse Events

Participants were prompted weekly to report any adverse events that occurred. AEs were summarized by group, severity, relationship to products, outcome and seriousness.

### Data Evaluability and Statistical Analysis

#### GSRS

Participants were considered evaluable for CLB101^TM^ analysis if they completed baseline and follow-up GSRS questionnaires during the Study Product/Placebo use period. Participants needed to have completed their baseline measurements in order to be included in any analysis of within-individual change.

The individual changes in the GSRS were aggregated from the baseline period to week 4 (end of product use). The pre-specified exploratory secondary outcome was change in GSRS composite from baseline to Week 4, evaluated by two-sided Mann-Whitney U test at a two-sided alpha of 0.05 without multiplicity correction. An identical analysis was conducted at Week 2 as an intermediate timepoint. GSRS subscales (Reflux, Abdominal Pain, Indigestion, Diarrhea, Constipation) were treated as an exploratory sub-family, with between-group change scores evaluated by Mann-Whitney U test and p-values adjusted using the Benjamini-Hochberg false discovery rate procedure (FDR-BH, k=5). Starred annotations on figures reflect the effective corrected p-value: uncorrected for the composite primary outcome, FDR-BH-adjusted for subscales.

The proportion of participants achieving improvement meeting or exceeding the published minimum clinically important difference (MCID) threshold was compared between groups at Weeks 2 and 4 using two-sided Fisher’s exact test. MCID thresholds applied were: Abdominal Pain 0.6, Reflux 0.8, Diarrhea 0.35, Indigestion 0.7, Constipation 0.7, and GSRS composite 3.13 points (Galafold, 2018). Improvement was defined as a decrease from baseline meeting or exceeding the metric-specific threshold.

#### Daily Survey Data

Within-group temporal trends were assessed using the Mann-Kendall trend test; between-group differences at the end-of-study window were evaluated by two-sided Mann-Whitney U test. A sensitivity analysis excluded participants who reported antibiotic use during the study period.

#### Intestinal Permeability: Dual Sugar Test

Two metrics were evaluated: lactulose percentage recovery (in which a decrease represents improvement), mannitol percentage recovery (in which an increase represents improvement), and the L/M ratio (in which a decrease represents improvement). Within-group pre-to-post changes were assessed by Wilcoxon signed-rank test; between-group differences in change scores by Mann-Whitney U test; and the proportion of participants improving was compared by Fisher’s exact test (FET). Outliers were defined as values more than 2 SD from the pooled mean across both timepoints and groups per metric and examined in sensitivity analyses.

#### Gut Microbiome, Butyrate Producer and Fungal biome Analysis

Change from baseline in gut microbiome and mycobiome composition at Week 4 was analyzed and measured for stool specimens between placebo and study product groups. Precomputed bacterial 16S and fungal ITS taxonomic count tables were matched to participant metadata. Primary longitudinal analyses included only participants with paired samples. Genus-level counts were prevalence-filtered, and the most abundant retained taxa were analyzed. A small pseudocount was added before applying the centered log-ratio (CLR) transformation. Within-group changes were evaluated using paired Wilcoxon signed-rank tests, while between-group differences in participant-level changes were evaluated using two-sided Wilcoxon rank-sum tests, equivalent to Mann–Whitney U tests.

Taxon-level p-values were adjusted separately within the 16S and ITS analyses using the Benjamini–Hochberg false-discovery-rate procedure, with adjusted values reported as q-values. In addition to the change from baseline in the abundance of *A. caccae* CLB101^TM^ at Week 4 was measured by qPCR in stool specimens and compared between placebo and study product groups.

#### Relative Abundance

As an exploratory analysis, relative abundance of *Faecalibacterium prausnitzii* was calculated for each sample as the number of reads assigned to *Faecalibacterium prausnitzii* divided by the total number of reads assigned to the domain Bacteria after quality control. Reads classified as Archaea or lacking a domain-level taxonomic assignment were excluded from the primary denominator.

#### Dietary and Product Intake

Univariate statistics were generated to describe the distribution of patient characteristics and outcome data. Dietary intake was assessed via a self-reported diary. Food category distributions were compared between groups at baseline and end of study using chi-square and Fisher’s exact tests. Product use compliance was tracked via daily check-in; adherence was defined as the proportion of days with confirmed product use. Participants with >= 80% compliance were classified as adherent.

#### Blood and Stool Composition

Other exploratory secondary objectives, including gastrointestinal symptoms, change from baseline in serum LPS-BP, ALT, and AST along with fecal calprotectin, zonulin, lactoferrin and butyrate were evaluated using both absolute before and after scores, and by comparing within-individual magnitude and direction of change. For a table with published reference ranges for each biomarker, see: **Supplemental Table 2**.

## RESULTS

### Recruitment and Conduct

This study was approved by Sterling Institutional Review Board (14583-NACraft) and registered with ClinicalTrials.gov (NCT07336615). All participants gave informed consent.

### Sample Size and Demographics

83 participants were randomized, and 12 were withdrawn due to: loss to follow-up (4), participant decision (3), and antibiotic use (5). N=71 participants were evaluated across all endpoints. Due to a technical software error GSRS was only captured at week 4 in (n=45). Separately, biomarker sampling n was dependent on sample sufficiency, ranging from 21 for the dual sugar test to 38 for most blood biomarkers, see **Figure 4** and **Table 3** for details. Demographics are described in **Table 2**. and did not differ significantly in any subcohort (GSRS or analyte sampling, data not shown). Cohort was 77.5% female, aged a mean (SD) of 42.1 (12), and had a BMI mean (SD) of 31.3 (3.6). Sample size for biomarker collection varied slightly by biomarker depending on sample quality, and sample sizes are indicated for each analysis in the corresponding figure (**Figure 4-5**. **Table 3**).

**Table 2.**
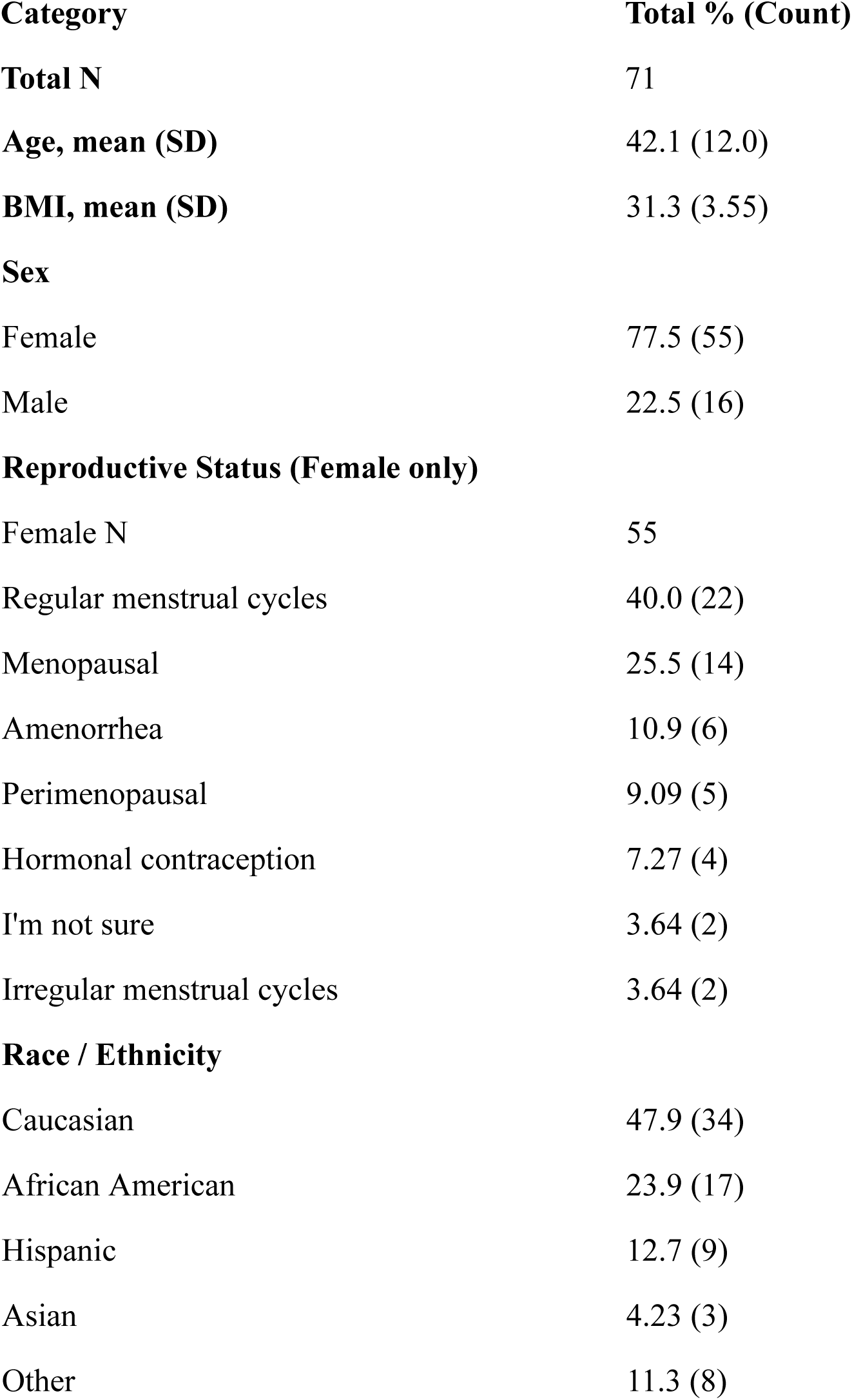
Demographics.

**Table 3.** Adverse Events.

| Group | Number of AEs | Description | Severity | Note |
| --- | --- | --- | --- | --- |
| CLB101™ | 1 | Stomach Jitters | Mild | Resolved |
| CLB101™ | 1 | Diarrhea | Severe | Resolved |
| CLB101™ | 1 | Nausea | Mild | Resolved |
| Placebo | 0 |  |  |  |

**Table 3.** Blood and Stool Biomarkers Indicate Absence of Intestinal Barrier Dysfunction.

| Biomarker | CLB101<br>n | CLB101 Mean<br>Change (SD) | Placebo<br>n | Placebo Mean<br>Change (SD) | p-value (Mann-Whitney U,<br>between groups) |
| --- | --- | --- | --- | --- | --- |
| Stool Butyrate | 21 | -5.28 (14.78) | 17 | +1.70 (13.80) | 0.022 |
| A. caccae<br>qPCR | 21 | -0.01 (0.82) | 17 | -0.09 (0.44) | 0.52 |
| Serum ALT | 21 | +3.50 (44.99) | 17 | +5.13 (15.56) | 0.161 |
| Serum AST | 21 | +3.05 (63.01) | 17 | +0.57 (25.90) | 0.659 |
| Serum LBP | 21 | -0.07 (1.35) | 17 | -0.12 (1.34) | 0.538 |
| Stool Calprotectin | 21 | -7.83 (109.22) | 17 | -23.65 (62.13) | 0.369 |
| Stool Lactoferrin | 21 | -2.92 (84.88) | 17 | -16.97 (56.53) | 0.953 |
| Stool Zonulin | 20 | -9.60 (108.15) | 17 | -7.04 (77.66) | 0.594 |
| Stool sIgA | 21 | +0.98 (249.42) | 17 | -32.04 (310.47) | 0.747 |

(Mann-Whitney U, uncorrected).

### Adverse Events

3 Adverse events were reported as possibly attributable to product use. These all occurred in CLB101^TM^ and consisted of mild stomach jitters, mild nausea, and 1 case of severe diarrhea. All resolved and participants opted to continue the study without further AE reports. The participant who experienced diarrhea noted that stools were of variable consistency throughout the study. For adverse events *unlikely* to be related to study product use, no AE differed in prevalence by group. Non-product related AEs (CLB101^TM^, n=3) and placebo (n=5), consisted of menstrual cramps, erectile dysfunction, temporal mandibular joint pain, colds and flu. See **Table 3**.

### Gastrointestinal Symptom Rating Scale (GSRS)

No baseline differences were observed in GSRS score or subscores. Total GSRS score (MWU p=0.04), reflux subscore (MWU FDR-BH p=0.02) and abdominal pain (MWU FDR-BH p=0.02) subscore decreased to a significantly greater degree in CLB101^TM^ than in placebo at week 4 (**Figure 2**). Subscores for indigestion, diarrhea, and constipation trended toward greater reduction with CLB101^TM^ supplementation, though these differences were not statistically significant. See **Supplementary Materials** for complete week 2 scores.

**Figure 1.**
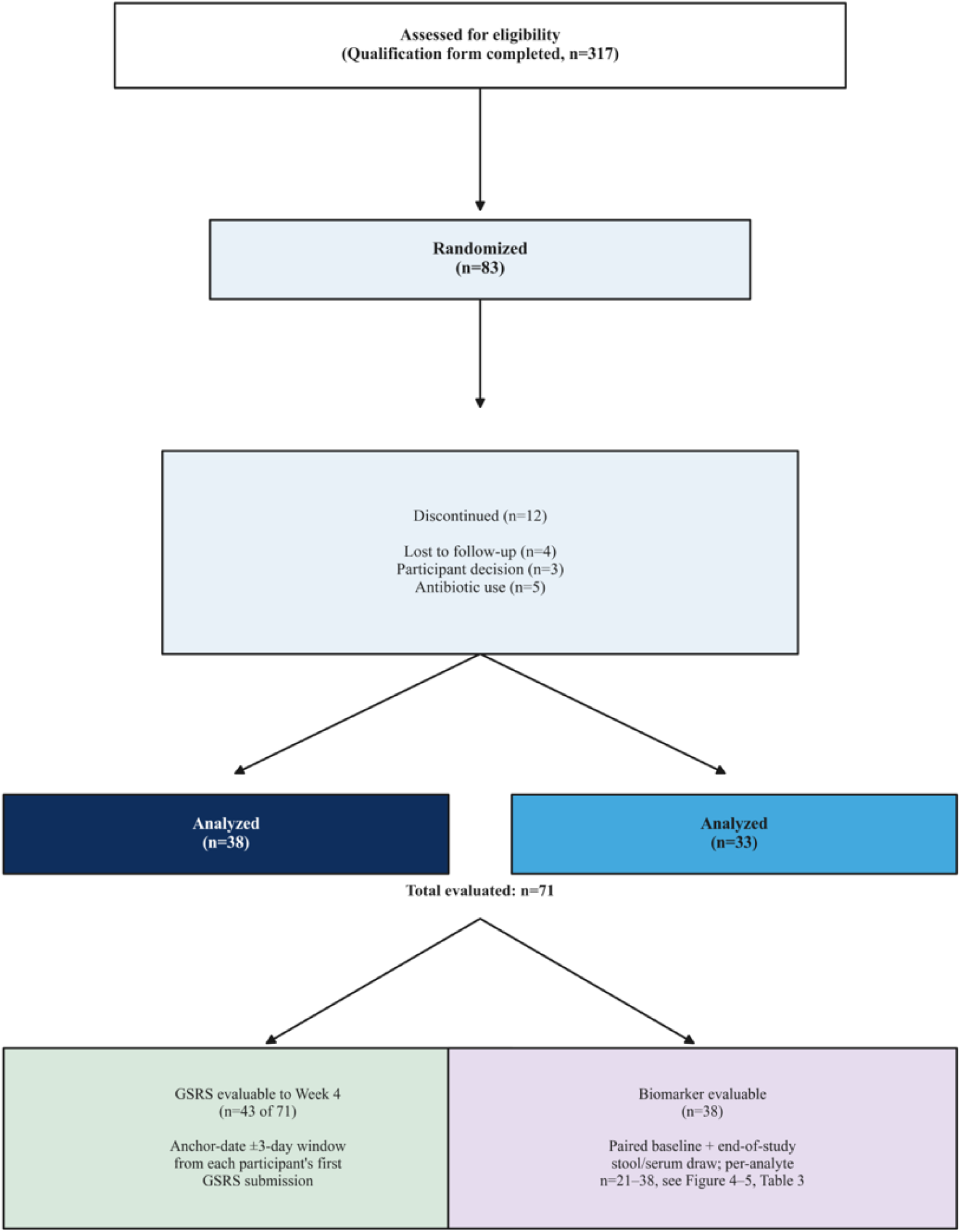
Consort Diagram.

**Figure 2.**
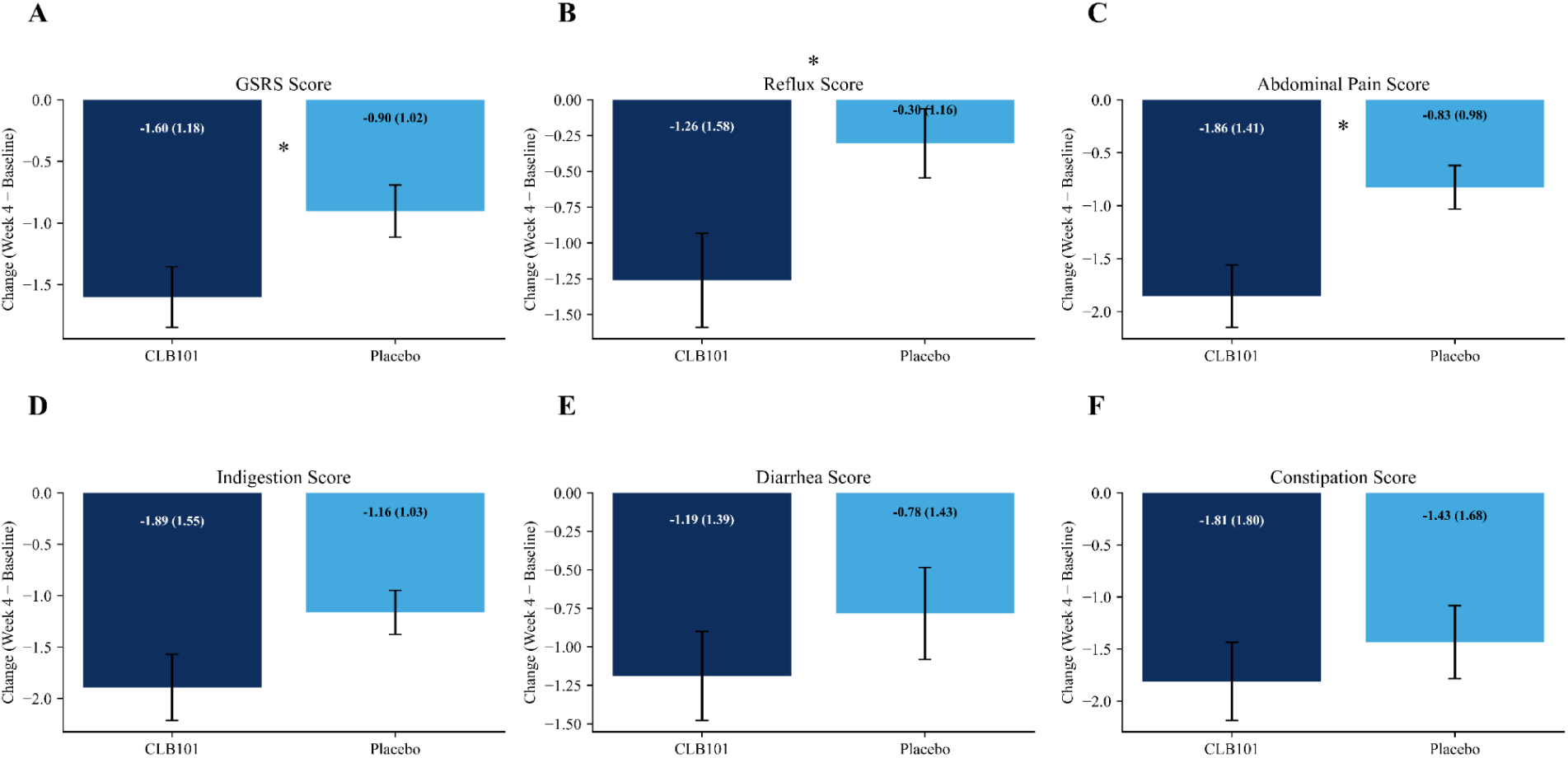
CLB101^TM^ Significantly Reduces GSRS Score, Reflux and Abdominal Pain Subscores After 4 Weeks. Bar charts of mean (SEM) of GSRS total (A) and sub-scores (B-F). * indicates statistical significance (p<0.05). CLB101^TM^ is shown in dark blue and placebo in light blue.

### Daily Digestive Symptoms Rating

Digestive symptoms of abdominal discomfort, bloating, flatulence, and quality of life trended toward improvement in both groups across the study period. Groups did not differ at baseline for any metric. CLB101^TM^ exhibited a non-statistically-significant trend toward greater reduction of abdominal discomfort, bloating and flatulence than placebo (p=0.06-0.09) **(Figure 3**). The number of daily bowel movements did not vary statistically across the study period in either group, with CLB101^TM^ mean ranging from 1.4 to 1.5 and placebo from 1.5 to 1.7. Stool form scale was rated at a 2.00 (0.67) in CLB101^TM^ and 1.95 (0.59) in placebo, corresponding to regular stool.

**Figure 3.**
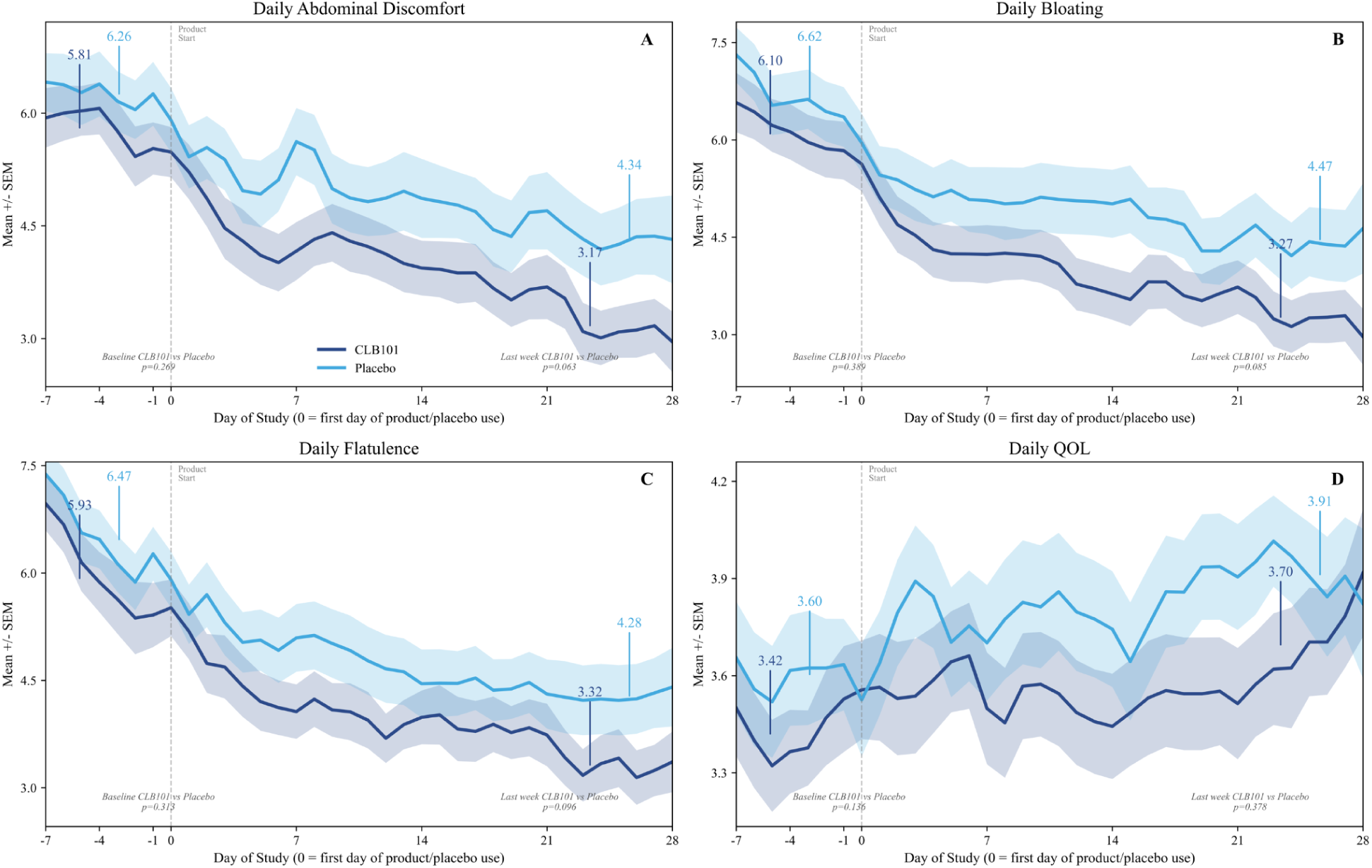
Daily Digestive Symptoms Decrease More Rapidly in CLB101^TM^. Mean with shaded SEM of Daily Abdominal Discomfort (A), Bloating (B), Flatulence (C), and Quality of Life (D). CLB101^TM^ is shown in dark blue and placebo in light blue.

### Intestinal Permeability Dual Sugar Test

Dual sugar absorption and ratio did not change significantly by timepoint or group (Figure 4). Mean change (EOS - Baseline) +/- SD: L/M ratio: CLB101^TM^ −0.028 +/- 0.120, placebo +0.013 +/- 0.050; Lactulose % recovery -- CLB101^TM^ +0.252 +/- 1.44, placebo +0.66 +/- 1.69; Mannitol % recovery -- CLB101^TM^ +3.36 +/- 18.2, placebo +1.70 +/- 6.29. Two participants with confirmed dietary mannitol contamination (EOS mannitol recovery >100%) were excluded from L/M ratio and mannitol analyses (**Figure 4** panels A, C, D, F).

**Figure 4.**
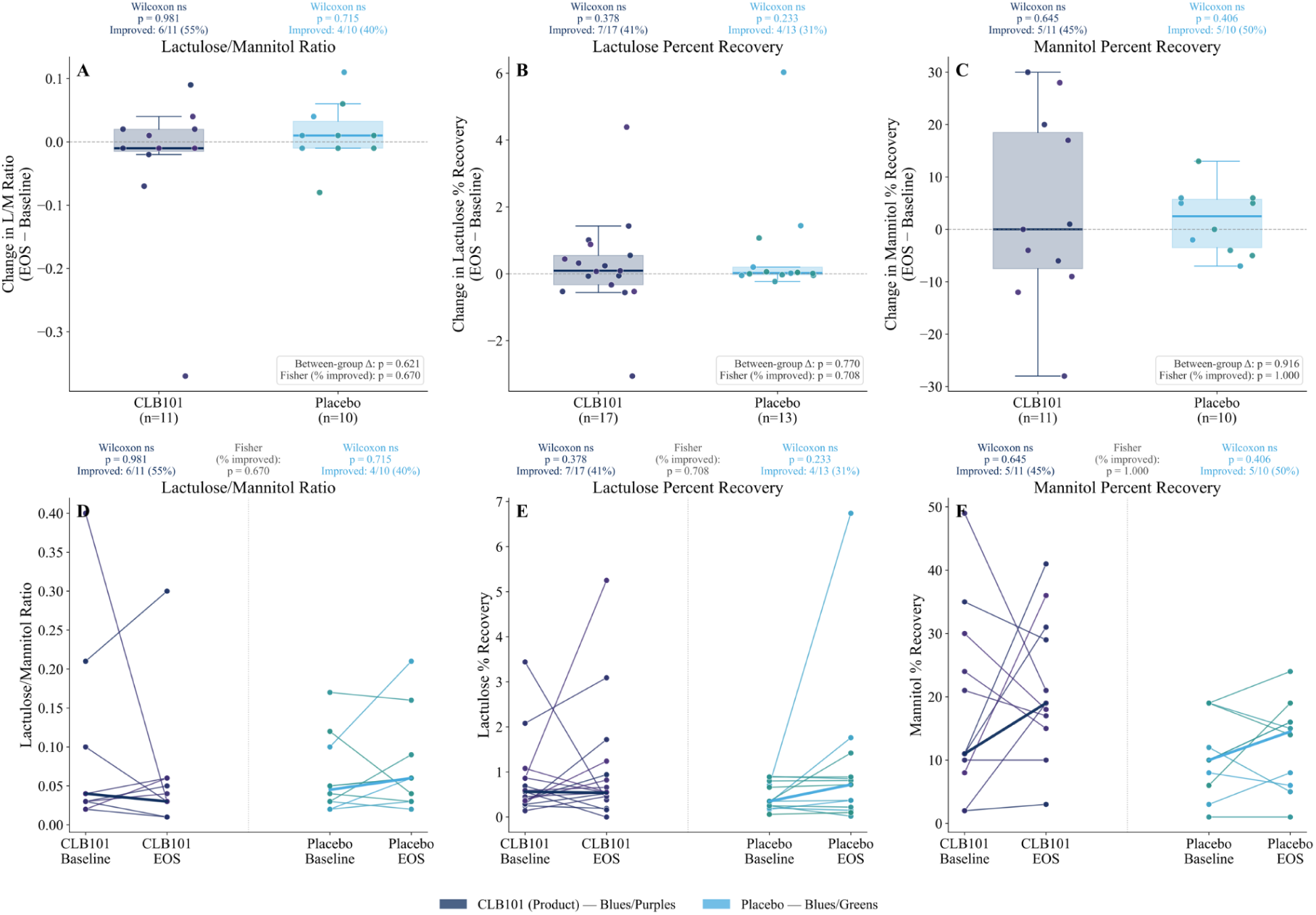
Dual Sugar Test Indicates Absence of Intestinal Barrier Dysfunction. Intestinal Permeability: Dual Sugar Test. Top row (A-C): box and whiskers with overlaid individuals of change from baseline to end of study (EOS) for lactulose/mannitol (L/M) ratio (A), lactulose percent recovery (B), and mannitol percent recovery (C). Bottom row (panels D-F): individual baseline-EOS trajectories for the same three metrics. Each line represents one participant; thick lines indicate group medians. Statistical annotations: Wilcoxon signed-rank test (within-group pre-post change); Mann-Whitney U (between-group difference in change scores); Fisher’s exact test (difference in proportion of improvers between groups). CLB101^TM^ = active product (navy/purple tones); placebo = placebo (blue/green tones). Outliers >2 SD from the pooled mean were excluded.

### Microbiome Composition

#### A caccae Abundance

No significant within-group, between-group, or baseline-adjusted difference was observed in *A. caccae* qPCR abundance (ANCOVA p=0.33-0.87 across modes; between-group raw-change p=0.32-0.62; between-group percent-change p=0.46-0.49). The proportion of participants with increasing abundance was numerically higher in CLB101^TM^ than placebo in all three modes (Raw: 28.6% vs 17.6%; IQR: 17.6% vs 7.1%; +/-2SD: 26.3% vs 7.1%), consistent with expected differential colonization in the active arm, but none of these differences reached significance (Fisher p=0.21-0.61, n=14-21 per group).

#### Butyrate-Producing Taxa and Species

Comparison of the 16s rRNA gene sequencing results to the genomic database enabled 343 species-level assignments, representing 16.7% of all bacteria identified. This list of species was compared to an extensive bioinformatic analysis of microbial genomes identifying hundreds of taxa with the genomic capacity to produce butyrate (Vital et al., 2014). The most prominent butyrate-producing species detected across all fecal samples in our study was *Faecalibacterium prausnitzii*, which was present in 100% of the samples tested and was the second most abundant species detected overall, behind only *Bacteroides uniformis*. In an exploratory analysis, supplementation with CLB101^TM^ trended toward an increase in the relative abundance of *F. prausnitzii*, whereas those receiving a placebo saw a slight reduction (MWU of Log2 fold change between group p=0.18, data not shown).

### Stool Calprotectin, Lactoferrin and Zonulin

Neither calprotectin nor lactoferrin showed a statistically significant within-group, between-group, or baseline-adjusted difference (Calprotectin ANCOVA: p=0.113; Lactoferrin ANCOVA: p=0.114), although the adjusted difference numerically favored CLB101^TM^. Groups were imbalanced in baseline severity: 66.7% of CLB101^TM^ vs 88.2% of placebo were within the normal range for calprotectin at baseline, and 38.1% of CLB101^TM^ vs 70.6% of placebo for lactoferrin. The CLB101^TM^ group entered the study with more participants starting out of range on both markers. CLB101^TM^ zonulin showed a numerical decrease (−9.6 ng/mL);, as well as placebo (−7.0 ng/mL) (ANCOVA p>0.05). Baseline in-range proportion was 45.0% for CLB101^TM^ vs 29.4% for placebo.

### Stool Butyrate

CLB101^TM^ showed a numerical decrease in mean change from baseline while placebo showed a numerical increase across methods of outlier handling (CLB101^TM^ −5.28 nmol [SD 14.78] vs placebo +1.70 nmol [SD 13.80], between-group Mann-Whitney p=0.022 **(See Figure 5**). The same pattern held on a percent-change basis, where the between-group difference was the most significant result in this panel (CLB101^TM^ −12.7% vs placebo +56.6%; p=0.006). However, baseline-adjusted ANCOVA did not reach significance in any mode (p=0.336). Neither baseline butyrate (Pearson r=-0.05, p=0.77; Spearman rho=0.02, p=0.88) nor change in butyrate (Pearson r=0.12, p=0.49; Spearman rho=0.07, p=0.67) was significantly correlated with GSRS change. N-values represent samples with enough recoverable mass for analysis.

**Figure 5.**
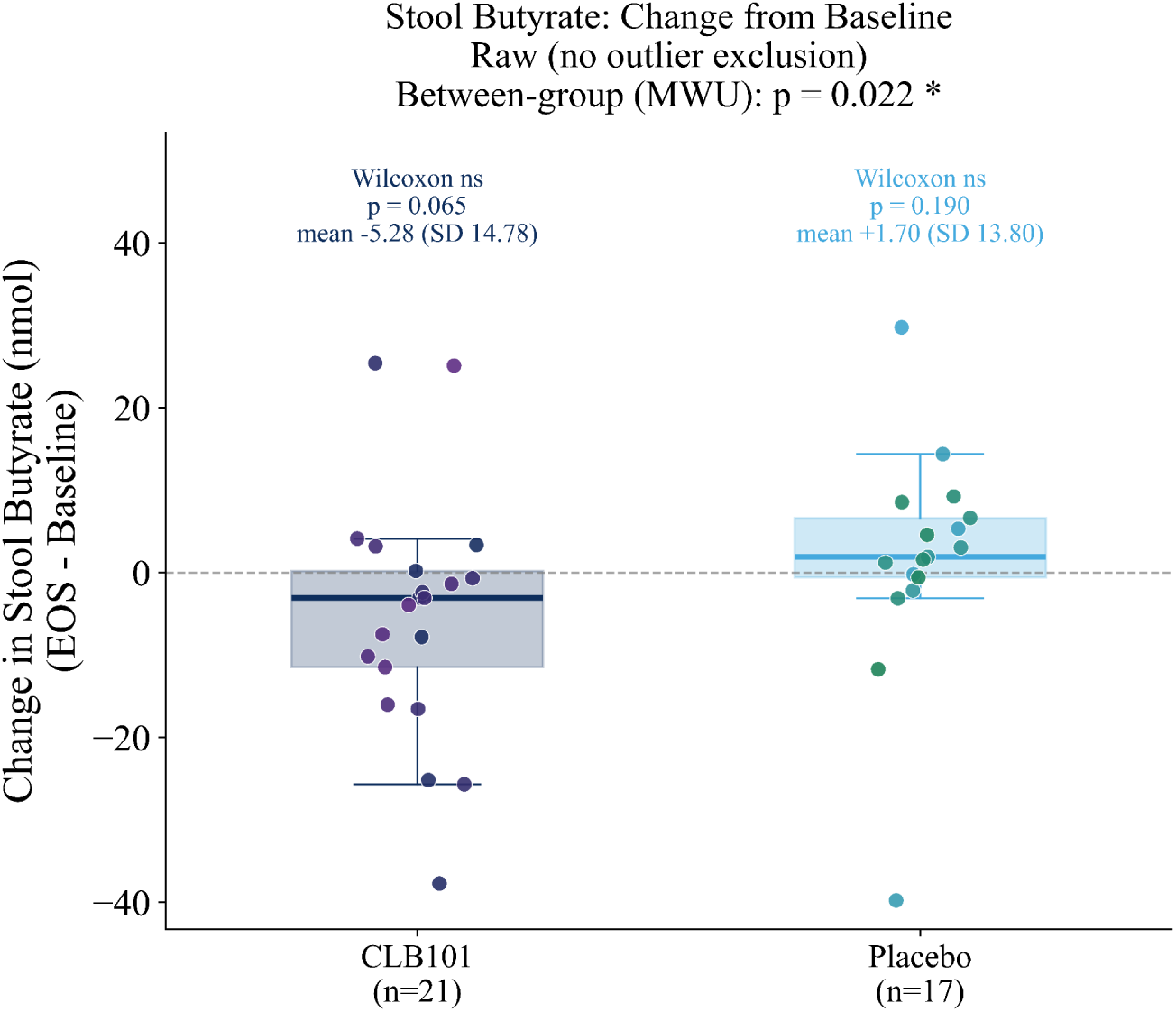
Excreted Butyrate is Significantly Reduced Following CLB101 Supplementation. Box and whisker plots with individual data scattered. CLB101^TM^ is shown in dark blue, and placebo in light blue.

### Stool sIgA

No significant within-group, between-group, or baseline-adjusted difference was observed (ANCOVA p=0.53 across modes). Using an in-range/out-of-range framing instead: 81.0% of CLB101^TM^ and 70.6% of placebo were within the literature reference range (510-2040 ug/g) at baseline. Among participants out of range at baseline, normalization rates by EOS were similar between groups (CLB101^TM^ 1/4, placebo 1/5; Fisher p=1.00), as were newly-out-of-range rates (CLB101^TM^ 1/17, placebo 2/12).

### Serum LBP

No baseline-adjusted ANCOVA or percent-change comparison reached significance (p>0.05). Baseline in-range proportions were similar between groups (28.6% CLB101^TM^, 23.5% placebo, against the 5-10 ug/mL reference range), and out-of-range-at-baseline normalization rates did not differ significantly between groups in any mode.

### Serum ALT and AST

AST showed no signal in any analysis (ANCOVA p>0.05; no within- or between-group raw- or percent-change comparison reached significance). Restricted to participants who began with abnormal (out-of-range) ALT, placebo showed a significant within-group raw-value increase (Wilcoxon p=0.019; mean +7.68 IU/L) while CLB101^TM^ did not (p=0.099; mean +4.49 IU/L), though the between-group comparison in this subset was not significant (p=0.133).

No evidence was found that treatment effect depends on baseline severity for any biomarker, though power to detect such an interaction was limited by small out-of-range subgroup sizes.

## Discussion and Conclusions

This double-blind, placebo-controlled trial demonstrated that CLB101^TM^ is safe and tolerable as a probiotic supplement. Strikingly, CLB101^TM^ also significantly decreased GSRS scores despite a study population that did not display signs of adverse intestinal barrier function. Emergence of effect over 4 weeks is consistent with an onset lag expected for butyrate-producing strains (Sato et al., 2008; Gilijamse et al., 2020). The magnitude of improvement in the active group exceeded MCID thresholds for Reflux (61% vs 23%) and Abdominal Pain (78% vs 64%) (Galafold, 2018) of CLB101^TM^ participants respectively, compared with the placebo arm. The reduction in reflux symptomatology is mechanistically plausible given butyrate’s known role in modulating enteric motility and esophageal mucosal integrity (Hamer et al., 2008). Daily diary data showed numerically greater reductions in abdominal discomfort (24% lower), bloating (33% lower), and flatulence (22% lower) in the CLB101^TM^ arm at end of study, providing directional corroboration of the GSRS findings across a complementary measurement approach. While CLB101 may support increased barrier function in other populations, its impact on GI symptoms suggests more generalized applications for improving intestinal health in the population as a whole.

Despite these consistent improvements in subjective gastrointestinal health, we did not observe corresponding changes in biomarkers of “leaky gut”. Preclinical models demonstrated a strong association between *A. caccae*, butyrate production, and intestinal barrier integrity. We therefore hypothesized that oral supplementation with CLB101^TM^ in humans would reduce adverse intestinal permeability and thereby relieve gastrointestinal symptoms potentially caused by “leaky gut”. We interrogated a collection of biomarkers, largely because there is no strong consensus as to which biomarker(s) define the condition (Camilleri, 2023). Fecal zonulin levels largely exceeded the normal range suggesting barrier dysfunction, though interpretation of this biomarker is complicated by the fact that commercial ELISA kits can detect other members of the zonulin family that are not demonstrably associated with intestinal barrier function (Camilleri, 2023). The two other biomarkers of potential barrier dysfunction, the lactulose/mannitol ratio and serum LPB, were well within normal ranges. Collectively, these data strongly suggest that the participants in this study experienced digestive symptoms despite healthy and functional intestinal barriers.

Two other biomarkers employed in this study assessed CLB101^TM^’s impact on the gut microbiome and its outputs. Results of the qPCR analysis failed to demonstrate an increase in *A. caccae* after supplementation with CLB101^TM^. This is somewhat surprising and may indicate that CLB101^TM^ did not survive the journey through the entirety of the gastrointestinal tract, though the strong impact on reflux suggests that its protective effects manifested in the proximal GI tract. Furthermore, it’s possible that CLB101^TM^ did not survive the fecal collection or freezing processes, or that its spores are recalcitrant to the DNA extraction protocol used in the study, both of which will be evaluated in future studies.

Finally, fecal butyrate levels were reduced in the study population receiving CLB101™, which would seem counterintuitive for a probiotic purported to produce butyrate. However, quantifying the impact of any intervention on intestinal butyrate levels is notoriously difficult without invasive sampling methods (Cummings & Macfarlane, 1991). It has been well established that fecal butyrate concentrations are not predictive of intestinal concentrations (Cummings & Macfarlane, 1991), and while the serum butyrate concentration may be indicative of intestinal absorption, it also represents a fraction of butyrate that was not metabolized by colonocytes and was therefore not supportive of intestinal barrier function. For those reasons, successful delivery of butyrate to colonocytes can only be assessed indirectly. Changes in the abundance of other butyrate producers is one indirect assessment of increased intestinal butyrate. Delivery of butyrate to the colonic epithelium promotes β-oxidation, which results in the rapid consumption of oxygen (Kelly et al., 2015). Such an environment supports expansion of anaerobic bacteria, some proportion of which are capable of further butyrate production (Wang et al., 2023). We therefore sought to determine if CLB101™-mediated butyrate delivery enriched populations of other strains capable of producing butyrate. The genes required for butyrate production do not track with phylogenetic groups, but instead are dispersed across numerous bacterial taxa, making it necessary to assess butyrogenic capacity at the species level. The trend toward CLB101^TM^-associated increase in *F. prausnitzii*, the most prominent butyrate producer identified in our study, suggests that CLB101^TM^ may have delivered butyrate to colonocytes, though a further interrogation of microbiome effects will require larger cohort sizes.

The study had several limitations. The primary outcome of the study was safety and tolerability, and exploratory biomarker trends will require future studies in larger sample sizes to confirm. Both groups showed large within-group symptom improvements consistent with regression to the mean in a symptomatic sample. Additionally, the 4 week treatment window may be insufficient for maximal colonization and butyrate-mediated effects, particularly in participants with established dysbiosis.

This study provides the first clinical evidence that CLB101^TM^ (*A. caccae*) is well-tolerated and produces significant improvements in overall GI symptom burden, reflux, and abdominal pain after four weeks of daily supplementation in overweight symptomatic adults. These findings are consistent with CLB101^TM^’s mechanism as a butyrate producer supporting intestinal homeostasis, and align with prior evidence for related butyrate-producing organisms (Gilijamse et al., 2020). The commercial availability of direct butyrate-producing strains at clinically significant doses has been extremely limited due to the challenges inherent in manufacturing strict anaerobes at scale. In this regard, CLB101™ is a true next-generation strain and this clinical data represents a major advance in exploring the potential of supplementing with strictly-anaerobic, commensal human strains. Adequately powered trials with longer treatment durations, microbiome outcome measures, and enrichment for participants with confirmed barrier dysfunction are warranted to fully characterize the benefits and potential of CLB101^TM^.

## Data Availability

Data are available upon reasonable request from the corresponding author.

## Acknowledgements

We would like to thank ER Chan for his assistance in interpretation of microbiome data.

## SUPPLEMENTAL MATERIALS

**Figure S1.**
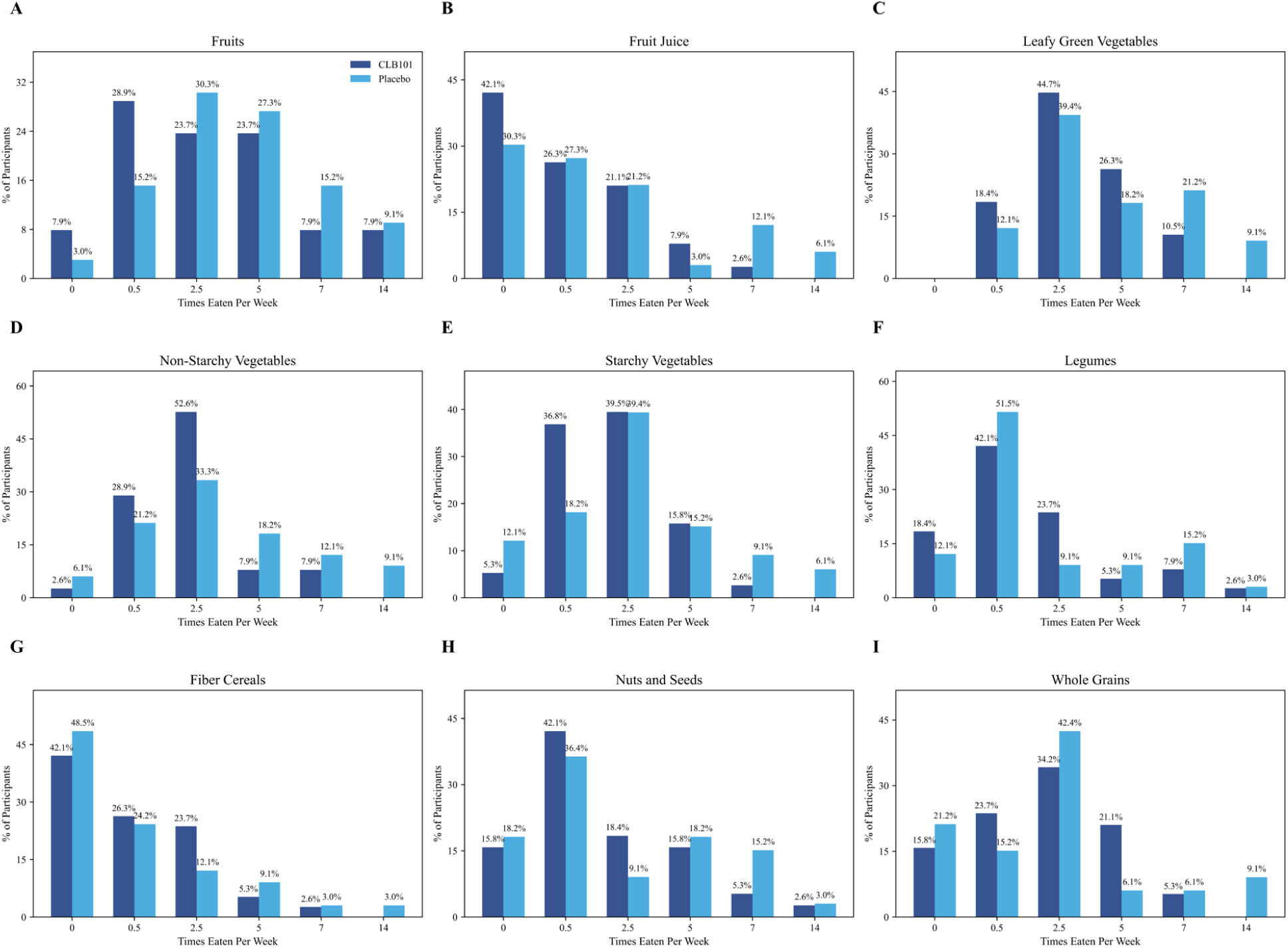
Dietary Style. Frequency of weekly consumption of key food groups reveals distribution did not vary by study group. CLB101^TM^ is shown in dark blue and placebo in light blue.

**Figure S3.**
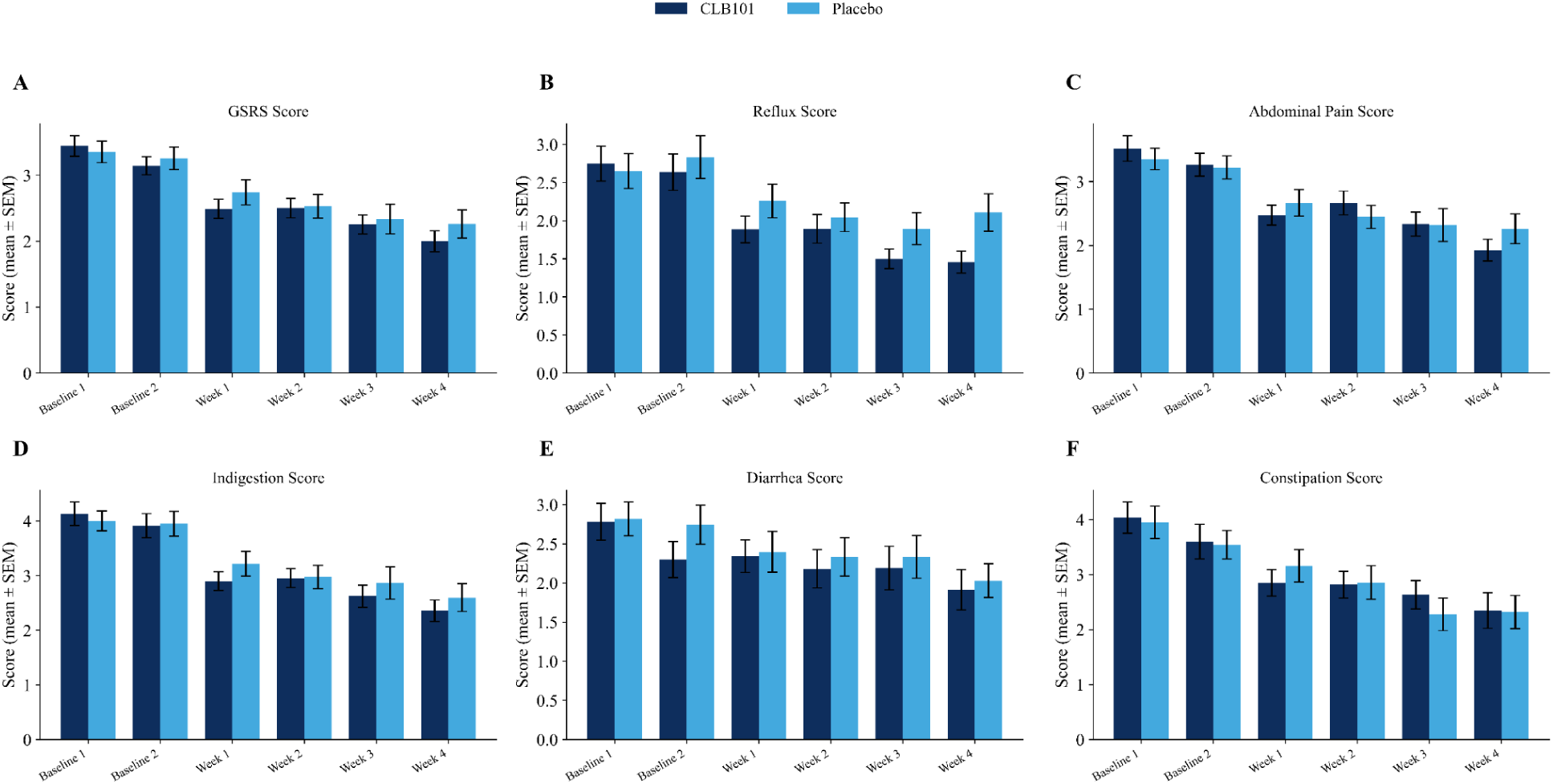
GSRS Score by Timepoint. GSRS by Timepoint GSRS Total Score (A) and sub score (B-F) means +/- SEM by time point. CLB101^TM^ is shown in dark blue and placebo in light blue.

**Figure S4.**
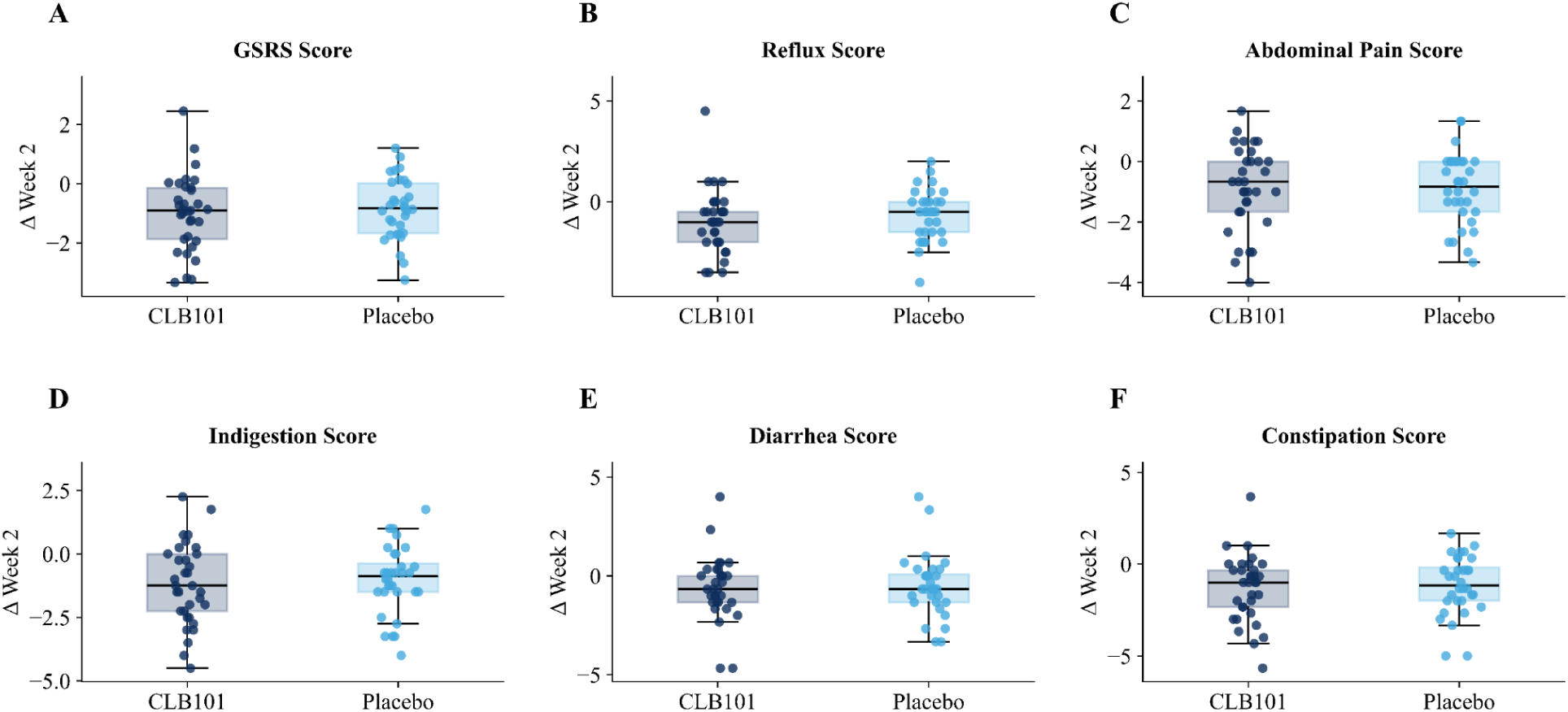
Week 2 Box and Whisker of Change in GSRS Scores to Week 2. Box and Whiskers of Change in GSRS Scores to Week 2 including total score (A) and subscores (B-F). CLB101^TM^ is shown in dark blue and placebo in light blue. Individual dots represent scattered individual changes from baseline to week 2.

**Figure S5.**
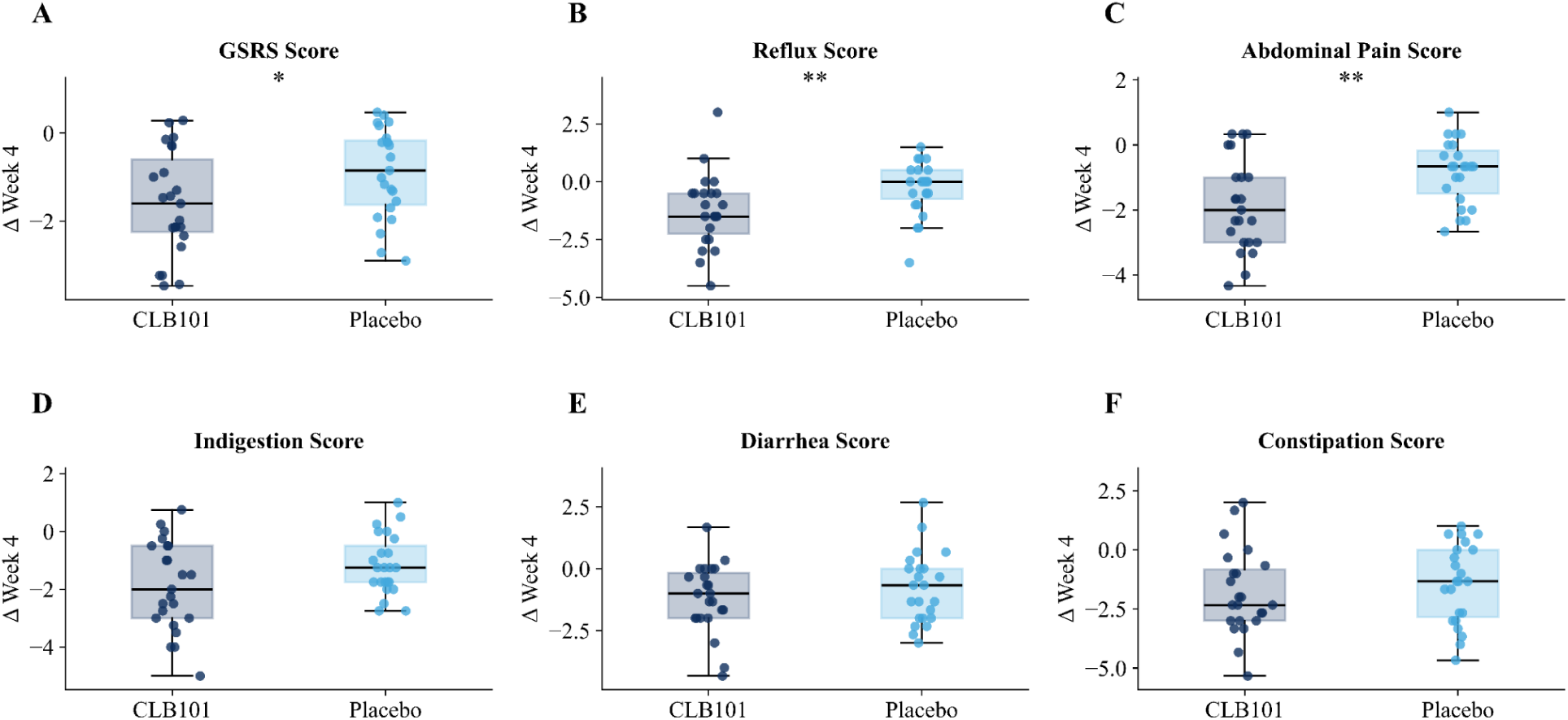
Week 4 Box and Whisker of Change in GSRS Scores. Box and Whiskers of Change in GSRS Scores to Week 4 including total score (A) and subscores (B-F). CLB101^TM^ is shown in dark blue and placebo in light blue. Individual dots represent scattered individual changes from baseline to week 2.

**Figure S6.**
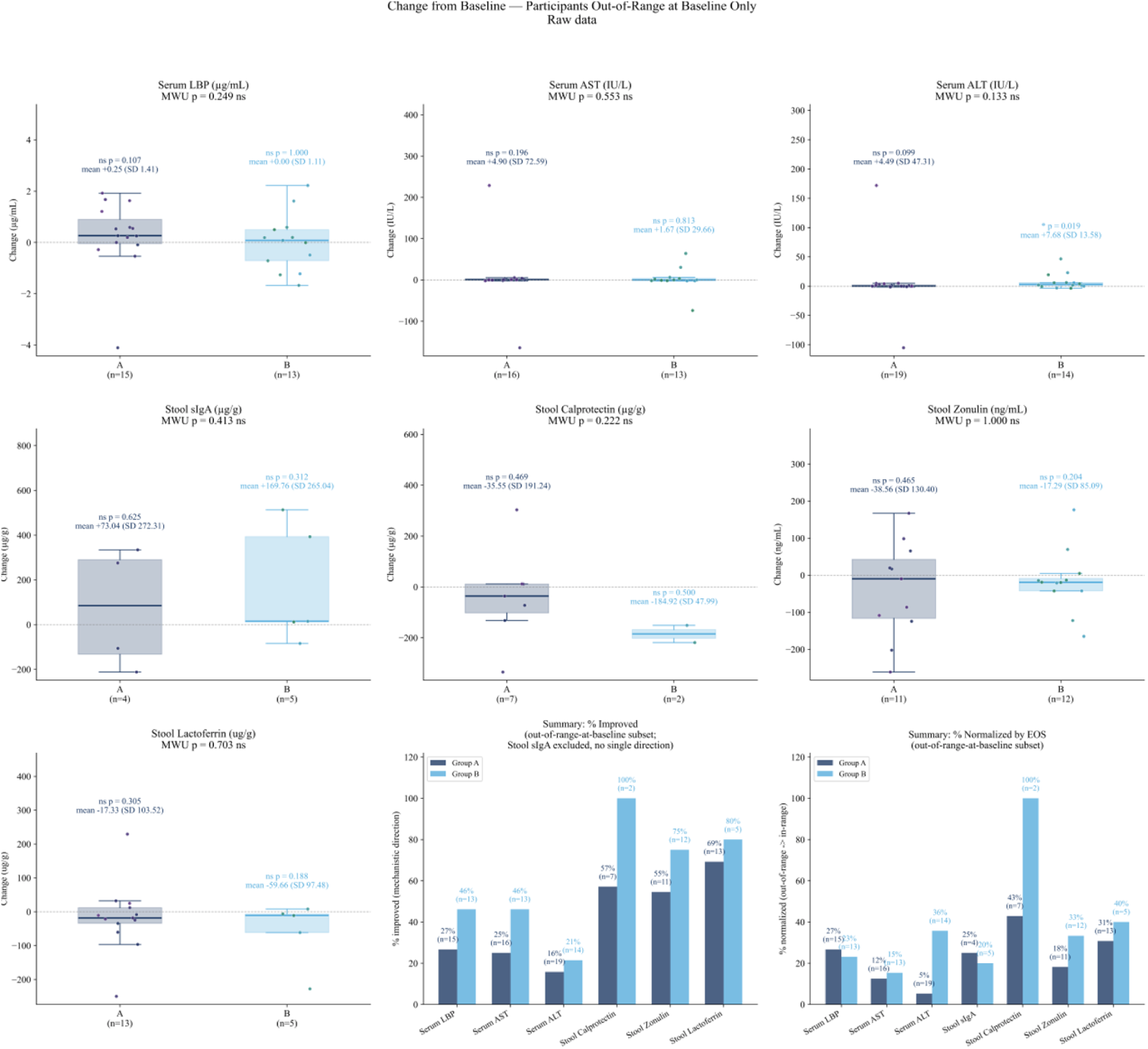
Analysis of Change in Biomarkers for Participants Who Were Out of Range at Baseline Only. **Box and Whiskers of Change in all Blood Biomarkers.** CLB101^TM^ is shown in dark blue and placebo in light blue. EOS indicates End of Study.

**Figure S7.**
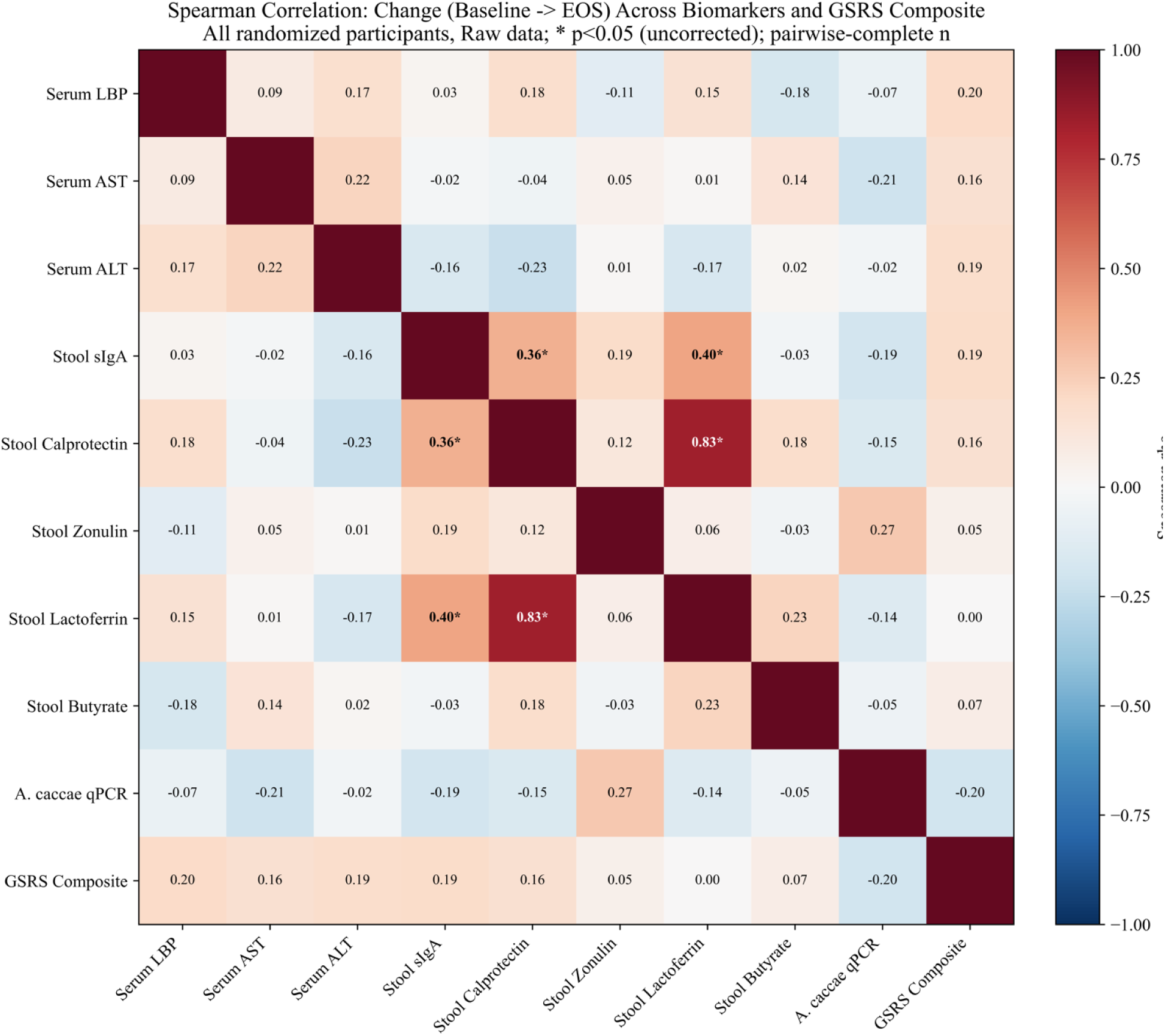
Spearman Correlation of Change from Baseline to End of Study in Stool and Blood Biomarkers, and GSRS Total Score. Color key indicates negative perfect correlation (dark blue), to positive perfect correlation (dark red). Data are amassed across arms.

**Table S1.** No Differences in Variability in Dietary Intake Across the Study Period.

| Absolute Change | CLB101<sup>TM</sup> % | placebo % | FDR p-value | FDR Reject |
| --- | --- | --- | --- | --- |
| 0 | 56.7 | 55.5 | 1 | No |
| 1 | 33.9 | 34.0 | 1 | No |
| 2 | 7.44 | 7.76 | 1 | No |
| 3 | 1.52 | 2.24 | 0.72 | No |
| 4 | 0.42 | 0.51 | 1 | No |
| 5 | 0 | 0.14 | 0.72 | No |

**Table S2.** Difference in % Improvers by GSRS Metric.

| **Metric** | **MCID Threshold** | **CLB101™ % improvers** | **placebo % improvers** | **Odds ratio** | **p** |
| --- | --- | --- | --- | --- | --- |
| Total Score | N/A | 17.4 | 0 | inf | 0.11 |
| **Reflux** | **0.8** | **60.97** | **22.7** | **5.29** | **0.02** |
| Abdominal Pain | 0.6 | 78.3 | 63.6 | 2.067 | 0.34 |
| Indigestion | 0.7 | 69.6 | 72.7 | 0.857 | 1 |
| Diarrhea | 0.35 | 65.2 | 54.5 | 1.56 | 0.54 |
| Constipation | 0.7 | 73.9 | 59.1 | 1.96 | 0.35 |

**Table S2.** Healthy Range Values for All Stool and Blood Analytes.

| Analyte | Healthy value |
| --- | --- |
| ALT | 7-55U/L (*Liver Function Tests - Mayo Clinic*, n.d.) |
| AST | 8-48U/L (*Liver Function Tests - Mayo Clinic*, n.d.) |
| Butyrate | 3.5-32.6μmol/g (McOrist et al., 2011) |
| Calprotectin | <50μg/g (Lamb & Mansfield, 2011) |
| Lactoferrin | <7.25μg/g (Lamb & Mansfield, 2011) |
| LBP | 5-10μg/mL (Gonzalez-Quintela et al., 2013) |
| sIgA | 510-2040μg/mL (Jendraszak et al., 2022) |
| Zonulin | <107ng/mL (Ciurea et al., 2025) |

**Table S3.** Baseline and End of Study Biomarker Values.

| Biomarker | CLB101<br>n | CLB101<br>Baseline<br>Mean<br>(SD) | CLB101<br>EOS<br>Mean<br>(SD) | Placebo<br>n | Placebo<br>Baseline<br>Mean<br>(SD) | Placebo<br>EOS<br>Mean<br>(SD) | Baseline<br>p-value<br>(Mann-Whitney<br>U,<br>between<br>groups) | EOS<br>p-value<br>(Mann-Whitney<br>U,<br>between<br>groups) | Baseline<br>FDR-p<br>(BH) | EOS<br>FDR-p<br>(BH) |
| --- | --- | --- | --- | --- | --- | --- | --- | --- | --- | --- |
| Stool<br>Butyrate | 21 | 18.58<br>(16.22) | 13.30<br>(12.18) | 17 | 13.42<br>(11.07) | 15.12<br>(11.84) | 0.463 | 0.538 | 0.993 | 0.836 |
| A. caccae<br>qPCR | 21 | 1.52<br>(0.88) | 1.50<br>(0.91) | 17 | 1.64<br>(1.14) | 1.55<br>(1.06) | 0.932 | 1.0 | 1.0 | 1.0 |
| Serum ALT | 21 | 7.31<br>(22.98) | 10.81<br>(37.30) | 17 | 4.65<br>(7.50) | 9.78<br>(13.03) | 0.705 | 0.071 | 0.993 | 0.213 |
| Serum AST | 21 | 12.26<br>(36.47) | 15.31<br>(49.30) | 17 | 9.49<br>(17.24) | 10.06<br>(17.37) | 0.393 | 0.965 | 0.993 | 1.0 |
| Serum LBP | 21 | 4.75<br>(1.55) | 4.69<br>(0.75) | 17 | 4.67<br>(0.93) | 4.56<br>(1.34) | 0.639 | 0.333 | 0.993 | 0.749 |
| Stool<br>Calprotecti<br>n | 21 | 58.85<br>(89.44) | 51.02<br>(91.29) | 17 | 33.29<br>(65.94) | 9.64<br>(6.75) | 0.115 | 0.01 | 0.603 | 0.09 |
| Stool<br>Lactoferrin | 21 | 46.71<br>(68.00) | 43.79<br>(71.99) | 17 | 27.69<br>(61.55) | 10.72<br>(19.15) | 0.134 | 0.069 | 0.603 | 0.213 |
| Stool<br>Zonulin | 20 | 144.78<br>(99.35) | 135.18<br>(89.80) | 17 | 149.79<br>(89.46) | 142.75<br>(94.55) | 0.772 | 0.988 | 0.993 | 1.0 |
| Stool sIgA | 21 | 781.95<br>(328.93) | 782.93<br>(361.37) | 17 | 749.37<br>(423.46) | 717.33<br>(355.23) | 1.0 | 0.557 | 1.0 | 0.836 |

*Values are Mean (SD) of raw measurements at Baseline and End-of-Study (EOS) among participants with both timepoints available (Raw mode, no outlier exclusion), matching the methodology of the Change-from-Baseline table. P-values are two-sided Mann-Whitney U tests comparing CLB101 vs. Placebo raw values at each timepoint independently (not a change score). FDR-p columns are Benjamini-Hochberg-corrected p-values, applied within each timepoint's family of 9 biomarker comparisons (Baseline and EOS corrected separately).*

## Supplemental Methods

### LPS-BP

Serum samples were diluted 1:100 (10 µL serum + 990 µL Assay Diluent B) and assayed in duplicate using a solid-phase sandwich ELISA. Standards (0.819–200 ng/mL, 7-point 2.5-fold serial dilution) were prepared in 1× Assay Diluent B. Following a 2.5-hour antigen binding incubation, biotinylated detection antibody was added for 1 hour, followed by Streptavidin-HRP for 45 minutes, with 4× washes between each step. TMB substrate was added for 30 minutes in the dark, and the reaction was stopped with Stop Solution. Absorbance was read at 450 nm. A 4-parameter logistic (4PL) curve was fit to standards in GraphPad Prism; sample concentrations were interpolated from the curve and multiplied by 100 to obtain true serum LBP in ng/mL.

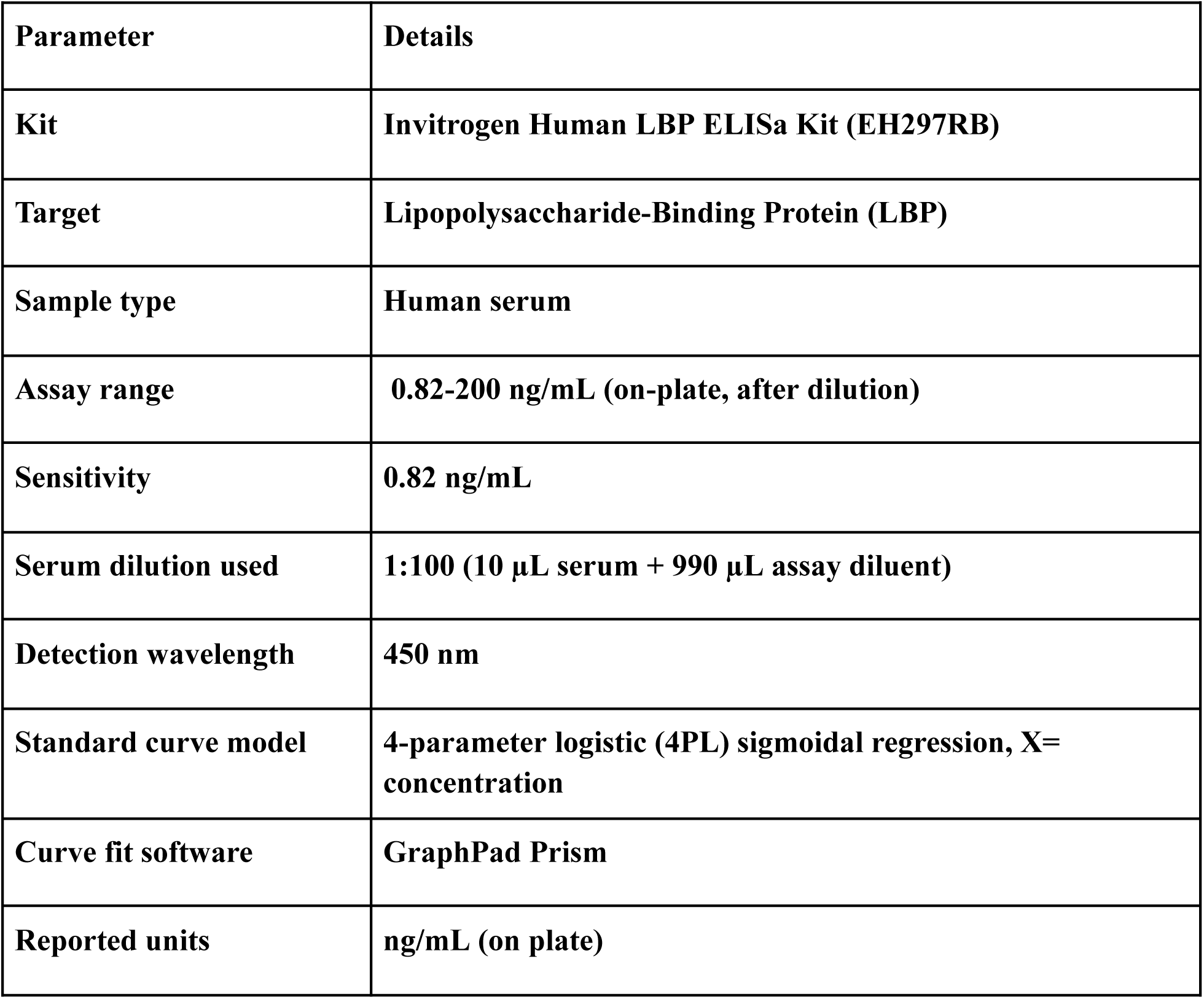

| Parameter | Details |
| --- | --- |
| **Kit** | **Invitrogen Human LBP ELISa Kit (EH297RB)** |
| **Target** | **Lipopolysaccharide-Binding Protein (LBP)** |
| **Sample type** | **Human serum** |
| **Assay range** | **0.82-200 ng/mL (on-plate, after dilution)** |
| **Sensitivity** | **0.82 ng/mL** |
| **Serum dilution used** | **1:100 (10 µL serum + 990 µL assay diluent)** |
| **Detection wavelength** | **450 nm** |
| **Standard curve model** | **4-parameter logistic (4PL) sigmoidal regression, X= concentration** |
| **Curve fit software** | **GraphPad Prism** |
| **Reported units** | **ng/mL (on plate)** |

### AST/ALT

For both AST & ALT, human serum was run undiluted (dilution factor = 1). Standards were prepared by adding fixed volumes of 2 mmol/L sodium pyruvate (0–8 µL) and substrate solution (12–20 µL) to each standard well with 5 µL buffer solution, creating a 5-point calibration series (0–190 Carmen units for AST; 0–200 Carmen units for ALT). After a 30-minute incubation at 37°C, 20 µL chromogenic agent (DNPH) was added to each well, followed by 5 µL of sample in the control wells, and incubated a further 20 minutes at 37°C. Then 200 µL of alkali reagent working solution was added and absorbance was read at 510 nm after 15 minutes at room temperature. A quadratic polynomial (y = ax² + bx + c) was fit to standards; enzyme activity (IU/L) was calculated as [a(ΔA₅₁₀)² + b(ΔA₅₁₀) + c] × 0.482 × df. In this case, df=1 as we did not do any dilution.

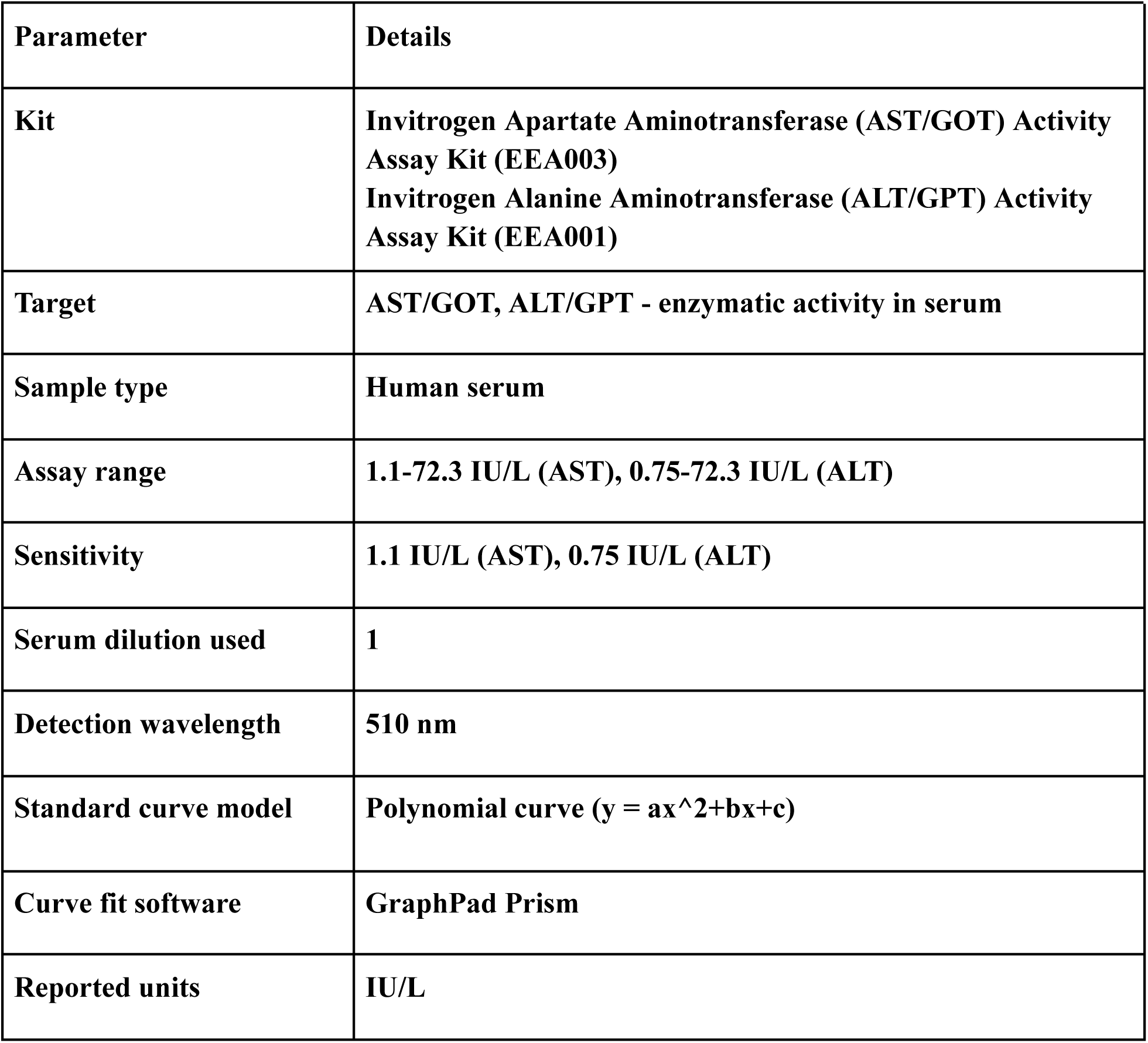

### sIGA

Fecal samples were extracted using the Stool Sample Application System (SAS): 15 mg stool was suspended in 1.5 mL IDK Extract® extraction buffer (1:2.5 diluted concentrate), yielding a 1:100 primary dilution (Dilution I). The supernatant was then further diluted 1:125 in the wash buffer (40 µL + 960 µL = 1:25; then 200 µL + 800 µL = 1:5), yielding a final dilution of 1:12,500. Samples were run in duplicate at 100 µL per well. Wells were pre-washed 5× with a wash buffer before use. Standards (0, 22.2, 66.6, 200, 600 ng/mL, lyophilized, reconstituted with 500 µL ultrapure water) and samples were incubated 1 hour at room temperature with shaking. After 5× washing, 100 µL diluted peroxidase-conjugate (mouse anti-sIgA, 1:101 in wash buffer) was added and incubated for 1 hour. After 5× washing, 100 µL TMB substrate was incubated 10–20 minutes in the dark, stopped with 100 µL stop solution, and absorbance read at 450 nm (reference 620 nm). A 4PL curve was fit in GraphPad Prism; interpolated values were multiplied by 12.5 (**2.5 in repeat run**) to obtain true fecal sIgA in µg/mL or µg/g.

| Parameter | Details |
| --- | --- |
| **Kit** | **Immundiagnostik AG IDK® sIgA ELISA (KR8870)** |
| **Target** | **Secretory IgA (sIgA)** |
| **Sample type** | **Stool extract (using IDK Extract)** |
| **Assay range** | **0 – 600 ng/mL (standards: 0, 22.2, 66.6, 200, 600 ng/mL)** |
| **Sensitivity (LoB)** | **2.088 ng/mL** |
| **Sample dilution used** | **1:12,500 (1:2,500 on rerun)** |
| **Detection wavelength** | **450 nm (reference: 620 nm or 690 nm)** |
| **Standard curve model** | **4-parameter logistic (4PL) sigmoidal regression, X = concentration** |
| **Curve fit software** | **GraphPad Prism** |
| **Reported units** | **ng/mL (converted to μg/g or μg/mL)** |

### Calprotectin

Fecal samples were extracted using the IDK Extract SAS (15 mg stool in 1.5 mL extraction buffer, 1:100). Extracts were further diluted 25× in dilution buffer (Range I: 16–625 µg/g). The assay used a single simultaneous incubation step: 50 µL of standard or diluted sample plus 50 µL of biotinylated capture antibody / HRP-conjugated detection antibody premix were loaded into streptavidin-coated wells and incubated 60 minutes at room temperature with shaking at 500 rpm. After 4× washing, 100 µL TMB substrate was added (up to 30 minutes), stopped with 100 µL oxalic acid stop solution, and absorbance read at 450 nm. A 4PL curve was fit in GraphPad Prism (log X-axis); interpolated values reported in µg/g.

| Parameter | Details |
| --- | --- |
| Kit | Hycult Biotech Human Fecal Calprotectin ELISA (HK382) |
| Target | Calprotectin (S100A8/A9 heterocomplex) |
| Sample type | Feces (stool extract using IDK Extract) |
| Assay range | 16–625 $\mu\text{g/g}$ feces (Range I, 50× dilution); 48–1,875 $\mu\text{g/g}$ (Range II, 150×) |
| Sensitivity | 16 $\mu\text{g/g}$ feces (minimum detectable) |
| Sample dilution used | 50× fecal extract in dilution buffer |
| Detection wavelength | 450 nm |
| Standard curve model | 4-parameter logistic (4PL) sigmoidal regression, X = concentration (log scale) |
| Curve fit software | GraphPad Prism |
| Reported units | $\mu\text{g/g}$ feces |

### Zonulin

Fecal samples were extracted using the Stool Sample Application System (SAS): 15 mg stool was suspended in 1.5 mL dilution buffer, yielding a 1:100 dilution. 150 µL of each stool extract, standard, or control was mixed with 150 µL of biotinylated ZFP tracer (1:101 diluted concentrate) in a separate tube immediately before use. Then 100 µL of each prepared mixture was loaded into anti-ZFP antibody-coated wells and incubated 1 hour with shaking at 350 rpm. After 5× washing, 100 µL streptavidin-peroxidase conjugate (1:101 diluted) was added for 1 hour. After 5× washing, 100 µL TMB substrate was incubated 10–20 minutes in the dark, stopped with 100 µL stop solution, and absorbance read at 450 nm (reference 620 nm). Because the assay is competitive, signal is inversely proportional to ZFP concentration. A 4PL curve was fit in GraphPad Prism; interpolated values were multiplied by 100 to obtain fecal ZFP in ng/mL.

| Parameter | Details |
| --- | --- |
| Kit | Immundiagnostik AG IDK® Zonulin ELISA (KR5600) |
| Target | Zonulin family peptides (ZFP) in stool |
| Sample type | Feces (stool extract) |
| Assay range | Per lot-specific specification data sheet (standards lyophilised; confirm concentrations from spec sheet) |
| Sensitivity (LoB) | 0.118 ng/mL (on-plate, before dilution correction) |
| Sample dilution used | 1:100 (15 mg stool + 1.5 mL dilution buffer via SAS) |
| Detection wavelength | 450 nm (reference: 620 nm or 690 nm); competitive ELISA — signal is INVERSE to concentration |
| Standard curve model | 4-parameter logistic (4PL) sigmoidal regression, X = concentration |
| Curve fit software | GraphPad Prism |
| Reported units | ng/mL (multiply interpolated value × 50) |

### Lactoferrin

Fecal samples were extracted in two steps: 15 mg stool was dissolved in 1.5 mL IDK Extract® extraction buffer (1:100; Dilution I), then 50 µL of Dilution I was added to 450 µL sample dilution buffer (1:10; Dilution II), yielding a final 1:1,000 dilution. Standards (lyophilized, concentrations per lot-specific spec sheet, reconstituted per spec sheet instructions) and samples were loaded at 100 µL per well and incubated 30 minutes at room temperature. After 5× washing, 100 µL peroxidase-conjugate (goat anti-human lactoferrin, 1:101 in wash buffer) was added and incubated 30 minutes. After 5× washing, 100 µL TMB substrate was incubated 10–20 minutes in the dark, stopped with 100 µL stop solution, and absorbance read at 450 nm (reference 620 nm). A 4PL curve was fit in GraphPad Prism; interpolated values were multiplied by 1,000 to obtain fecal lactoferrin in ng/mL. The kit reference value for normal stool is <7,200 ng/mL. If reporting in µg/g or µg/mL, divide by 1000.

| **Parameter** | **Details** |
| --- | --- |
| **Kit** | **Immundiagnostik AG Lactoferrin ELISA (K6870)** |
| **Target** | **Lactoferrin in stool (fecal marker of neutrophilic intestinal inflammation)** |
| **Sample type** | **Feces (stool extract)** |
| **Assay range** | **Per lot-specific specification data sheet; linear range 11.62–151.46 ng/mL (on-plate)** |
| **Sensitivity (LoB/LoD/LoQ)** | **LoB = 0.222 ng/mL; LoD = LoQ = 0.416 ng/mL** |
| **Sample dilution used** | **1:1000 total (15 mg stool + 1.5 mL IDK Extract® = 1:100; then 50 μL + 450 μL sample buffer = 1:10; final 1:1000)** |
| **Detection wavelength** | **450 nm (reference: 620 nm or 690 nm)** |
| **Standard curve model** | **4-parameter logistic (4PL) sigmoidal regression, X = concentration** |
| **Curve fit software** | **GraphPad Prism** |
| **Reported units** | **ng/mL (multiply interpolated value × 1000); divide by 1000 to report in μg/g or μg/mL** |

### Limit of Quantification Substitution

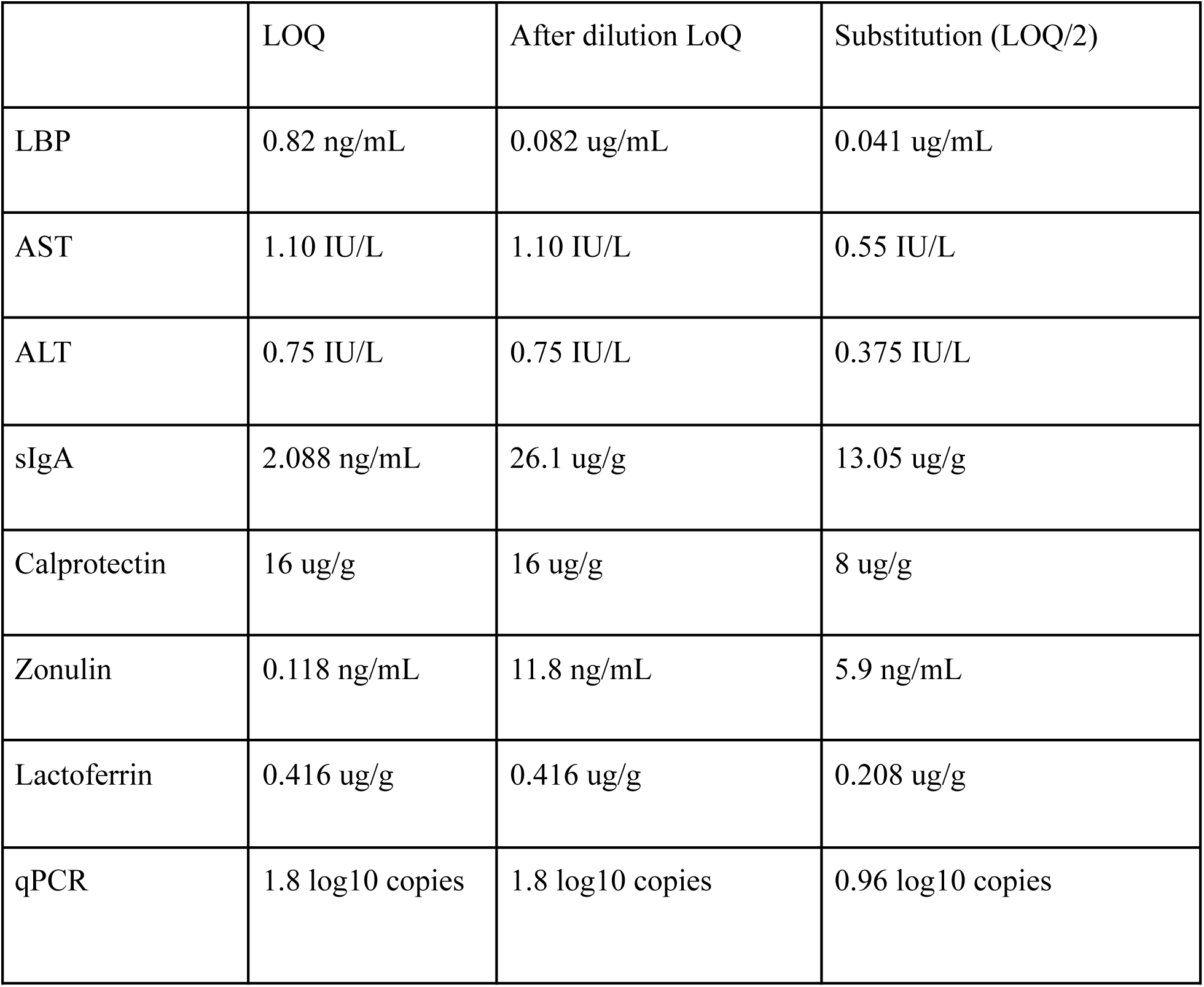

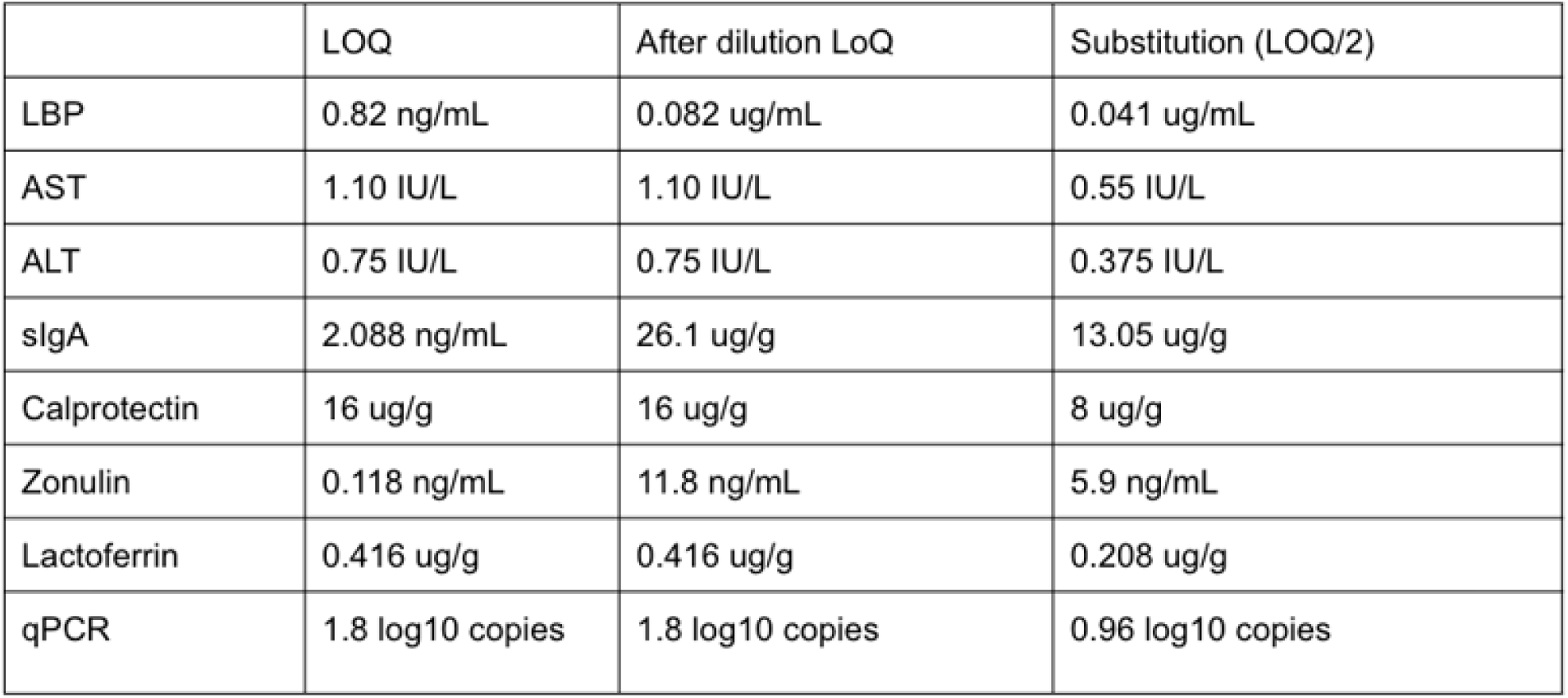

## REFERENCES

Belenguer, A., Duncan, S. H., Calder, A. G., Holtrop, G., Louis, P., Lobley, G. E., et al. 2006. Two routes of metabolic cross-feeding between Bifidobacterium adolescentis and butyrate-producing anaerobes from the human gut. Applied and Environmental Microbiology 72 (5): 3593–3599. doi: 10.1128/AEM.72.5.3593-3599.2006.

Brignardello, J., Morales, P., Diaz, E., Romero, J., Brunser, O., & Gotteland, M. 2010. Pilot study: alterations of intestinal microbiota in obese humans are not associated with colonic inflammation or disturbances of barrier function. Alimentary Pharmacology & Therapeutics 32 (11–12): 1307–1314. doi: 10.1111/j.1365-2036.2010.04475.x.

Camilleri, M. 2019. Leaky gut: mechanisms, measurement and clinical implications in humans. Gut 68 (8): 1516–1526. doi: 10.1136/gutjnl-2019-318427.

Camilleri, M. 2023. Is intestinal permeability increased in obesity? A review including the effects of dietary, pharmacological and surgical interventions on permeability and the microbiome. Diabetes, Obesity & Metabolism 25 (2): 325–330. doi: 10.1111/dom.14899.

Chia, L. W., Hornung, B. V. H., Aalvink, S., Schaap, P. J., de Vos, W. M., Knol, J., et al. 2018. Deciphering the trophic interaction between Akkermansia muciniphila and the butyrogenic gut commensal Anaerostipes caccae using a metatranscriptomic approach. Antonie Van Leeuwenhoek 111 (6): 859–873. doi: 10.1007/s10482-018-1040-x.

Chia, L. W., Mank, M., Blijenberg, B., Aalvink, S., Bongers, R. S., Stahl, B., et al. 2020a. Bacteroides thetaiotaomicron Fosters the Growth of Butyrate-Producing Anaerostipes caccae in the Presence of Lactose and Total Human Milk Carbohydrates. Microorganisms 8 (10): 1513. doi: 10.3390/microorganisms8101513.

Chia, L. W., Mank, M., Blijenberg, B., Aalvink, S., Bongers, R. S., Stahl, B., et al. 2020b. Bacteroides thetaiotaomicron Fosters the Growth of Butyrate-Producing Anaerostipes caccae in the Presence of Lactose and Total Human Milk Carbohydrates. Microorganisms 8 (10): 1513. doi: 10.3390/microorganisms8101513.

Chia, L. W., Mank, M., Blijenberg, B., Bongers, R. S., van Limpt, K., Wopereis, H., et al. 2021. Cross-feeding between Bifidobacterium infantis and Anaerostipes caccae on lactose and human milk oligosaccharides. Beneficial Microbes 12 (1): 69–83. doi: 10.3920/BM2020.0005.

Cummings, J. H. & Macfarlane, G. T. 1991. The control and consequences of bacterial fermentation in the human colon. The Journal of Applied Bacteriology 70 (6): 443–459. doi: 10.1111/j.1365-2672.1991.tb02739.x.

Dagbasi, A., Byrne, C., Blunt, D., Serrano-Contreras, J. I., Becker, G. F., Blanco, J. M., et al. 2024. Diet shapes the metabolite profile in the intact human ileum, which affects PYY release. Science Translational Medicine 16 (752): eadm8132. doi: 10.1126/scitranslmed.adm8132.

Damms-Machado, A., Louis, S., Schnitzer, A., Volynets, V., Rings, A., Basrai, M., et al. 2017. Gut permeability is related to body weight, fatty liver disease, and insulin resistance in obese individuals undergoing weight reduction. The American Journal of Clinical Nutrition 105 (1): 127–135. doi: 10.3945/ajcn.116.131110.

Duncan, S. H., Louis, P., & Flint, H. J. 2004. Lactate-utilizing bacteria, isolated from human feces, that produce butyrate as a major fermentation product. Applied and Environmental Microbiology 70 (10): 5810–5817. doi: 10.1128/AEM.70.10.5810-5817.2004.

Falony, G., Joossens, M., Vieira-Silva, S., Wang, J., Darzi, Y., Faust, K., et al. 2016. Population-level analysis of gut microbiome variation. Science 352 (6285): 560–564. doi: 10.1126/science.aad3503.

Feehley, T., Plunkett, C. H., Bao, R., Choi Hong, S. M., Culleen, E., Belda-Ferre, P., et al. 2019. Healthy infants harbor intestinal bacteria that protect against food allergy. Nature Medicine 25 (3): 448–453. doi: 10.1038/s41591-018-0324-z.

Galafold, M. 2018. Validity of Outcome Measures. Clinical Review Report: Migalastat (Galafold): (Amicus Therapeutics): Indication: Fabry Disease [Internet].

Gilijamse, P. W., Hartstra, A. V., Levin, E., Wortelboer, K., Serlie, M. J., Ackermans, M. T., et al. 2020. Treatment with Anaerobutyricum soehngenii: a pilot study of safety and dose–response effects on glucose metabolism in human subjects with metabolic syndrome. NPJ Biofilms and Microbiomes 6: 16. doi: 10.1038/s41522-020-0127-0.

Hamer, H. M., Jonkers, D., Venema, K., Vanhoutvin, S., Troost, F. J., & Brummer, R.-J. 2008. Review article: the role of butyrate on colonic function. Alimentary Pharmacology & Therapeutics 27 (2): 104–119. doi: 10.1111/j.1365-2036.2007.03562.x.

Hesser, L. A., Puente, A. A., Arnold, J., Ionescu, E., Mirmira, A., Talasani, N., et al. 2024. A synbiotic of Anaerostipes caccae and lactulose prevents and treats food allergy in mice. Cell Host & Microbe 32 (7): 1163–1176.e6. doi: 10.1016/j.chom.2024.05.019.

Hoshiko, H., Zeinstra, G. G., Lenaerts, K., Oosterink, E., Ariens, R. M. C., Mes, J. J., et al. 2021. An Observational Study to Evaluate the Association between Intestinal Permeability, Leaky Gut Related Markers, and Metabolic Health in Healthy Adults. Healthcare 9 (11): 1583. doi: 10.3390/healthcare9111583.

Kadowaki, R., Tanno, H., Maeno, S., & Endo, A. 2023. Spore-forming properties and enhanced oxygen tolerance of butyrate-producing Anaerostipes spp. Anaerobe 82: 102752. doi: 10.1016/j.anaerobe.2023.102752.

Lee, J.-Y., Tiffany, C. R., Mahan, S. P., Kellom, M., Rogers, A. W. L., Nguyen, H., et al. 2024. High fat intake sustains sorbitol intolerance after antibiotic-mediated Clostridia depletion from the gut microbiota. Cell 187 (5): 1191–1205.e15. doi: 10.1016/j.cell.2024.01.029.

Liu, H., Wang, J., He, T., Becker, S., Zhang, G., Li, D., et al. 2018. Butyrate: A Double-Edged Sword for Health? Advances in Nutrition 9 (1): 21–29. doi: 10.1093/advances/nmx009.

Lo Presti, A., Zorzi, F., Del Chierico, F., Altomare, A., Cocca, S., Avola, A., et al. 2019. Fecal and Mucosal Microbiota Profiling in Irritable Bowel Syndrome and Inflammatory Bowel Disease. Frontiers in Microbiology 10. doi: 10.3389/fmicb.2019.01655.

Modica, V., Glávits, R., Clewell, A., Endres, J. R., Hirka, G., Vértesi, A., et al. 2025. A Comprehensive Toxicological Safety Evaluation of Anaerostipes caccae. International Journal of Toxicology 44 (6): 471–487. doi: 10.1177/10915818251367353.

Peng, L., Li, Z.-R., Green, R. S., Holzman, I. R., & Lin, J. 2009. Butyrate enhances the intestinal barrier by facilitating tight junction assembly via activation of AMP-activated protein kinase in Caco-2 cell monolayers. The Journal of Nutrition 139 (9): 1619–1625. doi: 10.3945/jn.109.104638.

Qiao, Y. & Gao, X. 2026. Investigating the Interplay between the Gut Microbiota and Host Immunity in Gastroenteric Disorders: The Potential of Combined Drug Therapies to Restore Microbial-immune Homeostasis. Iranian Journal of Allergy, Asthma, and Immunology 25 (1): 32–44. doi: 10.18502/ijaai.v25i1.20434.

Recharla, N., Geesala, R., & Shi, X.-Z. 2023. Gut Microbial Metabolite Butyrate and Its Therapeutic Role in Inflammatory Bowel Disease: A Literature Review. Nutrients 15 (10): 2275. doi: 10.3390/nu15102275.

Ryan, F. J., Ahern, A. M., Fitzgerald, R. S., Laserna-Mendieta, E. J., Power, E. M., Clooney, A. G., et al. 2020. Colonic microbiota is associated with inflammation and host epigenomic alterations in inflammatory bowel disease. Nature Communications 11 (1): 1512. doi: 10.1038/s41467-020-15342-5.

Sato, T., Matsumoto, K., Okumura, T., Yokoi, W., Naito, E., Yoshida, Y., et al. 2008. Isolation of lactate-utilizing butyrate-producing bacteria from human feces and in vivo administration of Anaerostipes caccae strain L2 and galacto-oligosaccharides in a rat model. FEMS microbiology ecology 66 (3): 528–536. doi: 10.1111/j.1574-6941.2008.00528.x.

Schwiertz, A., Hold, G. L., Duncan, S. H., Gruhl, B., Collins, M. D., Lawson, P. A., et al. 2002. Anaerostipes caccae gen. nov., sp. nov., a new saccharolytic, acetate-utilising, butyrate-producing bacterium from human faeces. Systematic and Applied Microbiology 25 (1): 46–51. doi: 10.1078/0723-2020-00096.

Sola, L., Candeliere, F., Busi, E., Raimondi, S., Amaretti, A., & Rossi, M. 2026. A genomic atlas of gut clostridia: phylogeny, butyrate, and propionate production. Frontiers in Microbiology 17. doi: 10.3389/fmicb.2026.1761627.

Song, C.-H., Kim, N., Nam, R. H., Choi, S. I., Jang, J. Y., Kim, E. H., et al. 2024. The Possible Preventative Role of Lactate- and Butyrate-Producing Bacteria in Colorectal Carcinogenesis. Gut and Liver 18 (4): 654–666. doi: 10.5009/gnl230385.

Talley, N. J., Fullerton, S., Junghard, O., & Wiklund, I. 2001. Quality of life in patients with endoscopy-negative heartburn: reliability and sensitivity of disease-specific instruments. The American Journal of Gastroenterology 96 (7): 1998–2004. doi: 10.1016/S0002-9270(01)02495-9.

Teixeira, T. F. S., Souza, N. C. S., Chiarello, P. G., Franceschini, S. C. C., Bressan, J., Ferreira, C. L. L. F., et al. 2012. Intestinal permeability parameters in obese patients are correlated with metabolic syndrome risk factors. Clinical Nutrition 31 (5): 735–740. doi: 10.1016/j.clnu.2012.02.009.

